# Optimal Allocation of Limited Test Resources for the Quantification of COVID-19 Infections

**DOI:** 10.1101/2020.11.09.20228320

**Authors:** Michail Chatzimanolakis, Pascal Weber, George Arampatzis, Daniel Wälchli, Ivica Kičić, Petr Karnakov, Costas Papadimitriou, Petros Koumoutsakos

## Abstract

The systematic identification of infected individuals is critical for the containment of the COVID-19 pandemic. Presently, the spread of the disease is mostly quantified by the reported numbers of infections, hospitalizations, recoveries and deaths; these quantities inform epidemiology models that provide forecasts for the spread of the epidemic and guide policy making. The veracity of these forecasts depends on the discrepancy between the numbers of reported and unreported, yet infectious, individuals.

We combine Bayesian experimental design with an epidemiology model and propose a methodology for the optimal allocation of limited testing resources in space and time, which maximizes the information gain for such unreported infections. The proposed approach is applicable at the onset and spreading of the epidemic and can forewarn for a possible recurrence of the disease after relaxation of interventions. We examine its application in Switzerland; the open source software is, however, readily adaptable to countries around the world.

We find that following the proposed methodology can lead to vastly less uncertain predictions for the spread of the disease. Estimates of the effective reproduction number and of the future number of unreported infections are improved, which in turn can provide timely and systematic guidance for the effective identification of infectious individuals and for decision-making.

## 1. Introduction

The identification of unreported individuals infected by the SARS-CoV-2 virus is critical for the quantification, forecasting and planning of interventions during the COVID-19 pandemic [1]. Presently the spread of the disease is mostly quantified by the reported numbers of infections, hospitalizations, recoveries and deaths [2]. These quantities inform epidemiology models that provide short term forecasts for the spread of the epidemic, help quantify the role of possible interventions and guide policy making. The veracity of these forecasts depends on the discrepancy between the numbers of reported and unreported, yet infectious, individuals.

In recent months the estimation of unreported infections has been the subject of several testing campaigns [3, 4]. While there is valuable information being gathered, their estimates rely on testing individuals that are either already symptomatic or have been selected based on certain criteria (hospital visits, airport arrivals, geographic vicinity to researchers, etc.). Generic, randomized tests of the population are broadly applied but they have been hampered either by delays [5] or by insufficient numbers of test kits [6]. There is broad recognition that efficient testing strategies are critical for the timely identification of infectious individuals and the optimal allocation of resources [7]. However, targeted testing entails bias while randomized tests require access to a high percentage of the population with commensurate high costs. The quality of the data as well as the ways they are incorporated in the epidemiology models is critical for their predictions and for estimating their uncertainties [8]. Having the capability to minimize these uncertainties by suitably distributing in space and time a given number of test kits is the subject of this work. This optimal allocation of testing resources and the respective increase of the fidelity of forecasting models are essential to effective policy making throughout the pandemic.

Here, we present a methodology for the OPtimal Allocation of LImited Testing resourceS (OPALITS) that maximizes the information gain over any prior knowledge regarding infections. The method relies on forecasts by epidemiological models with parameters adjusted through Bayesian inference as data become available through suitable surveys [9]. The forecasts are combined with Bayesian experimental design [10, 11, 12] to determine the optimal test allocation in space and time for various objectives (minimize prediction uncertainty, maximize information gain of unreported infections). We emphasize that the proposed OPALITS is applicable in all stages of the pandemic, regardless of the availability of data.

We employ the *SEI*^*r*^*I*^*u*^*R* model [13] that quantifies the spread of a disease in a country’s population distributed in a number of communities that are interacting through mobility networks. The *SEI*^*r*^*I*^*u*^*R* model predicts the number of susceptible (*S*), exposed (*E*), infectious reported (*I*^*r*^), unreported (*I*^*u*^), and removed (*R*) individuals from the population. Here we focus on Switzerland and consider its cantons as the respective communities. The model parameters are: the relative transmission rate between reported and unreported infectious individuals (*µ*), the virus latency period (*Z*), the infectious period (*D*) and the reporting rate (*α*). The transmission rate (*β*) and the mobility factor (*θ*) are considered to be time dependent in order to account for government interventions. For all stages of the epidemic, the respective uncertainties of the model parameters are quantified and propagated using Bayesian inference. At the onset of the epidemic, the uncertainty is quantified through prior probability distributions. As data of daily infections become available, the uncertainty in model parameters is updated through Bayesian inference. The parameter probability distributions are used to propagate uncertainties in the model forecasts and can assist decision makers in quantifying risks associated with the progression of the disease. The proper quantification of uncertainty bounds in the model parameters has a profound effect on predictions of the disease dynamics [8]. Large uncertainty bounds around the most probable parameter values hinder the decision process for identifying effective interventions.

The OPALITS aims to assign limited test-kit resources to acquire data that would reduce the model prediction uncertainties. Minimizing the uncertainty of the model parameters leads to more reliable predictions for quantities such as the reproduction number [14]. Moreover, the reduced model uncertainties help minimize risks associated with the decision making process including timing, extent of interventions and probability of exceeding hospital capacity.

We quantify the information gain from these tests using a utility function [15, 12] based on the Kullback-Leibler divergence between the inferred posterior distribution and the current prior distribution of the model parameters. The prior can be formulated using the posterior distribution estimated from daily data of the infectious reported individuals up to the current date (see Materials and Methods). Hence, at any stage of the epidemic, the OPALITS provides guidance for the time and location/community where testing needs to be carried out to maximize the expected information gain regarding infections in a population.

We demonstrate the simplicity and applicability of the present method in estimating the spread of the coronavirus disease in the cantons of Switzerland. We find that the OPALITS methodology outperforms non-specific, randomized testing of sub-populations throughout the COVID-19 pandemic. The proposed strategy is readily applicable to other countries and the employed open source software can readily accommodate different epidemiological models.

## 2. Results

### Optimal Allocation of Limited Testing Resources (OPALITS) during the COVID-19 pandemic

We present the optimal test-kit allocation strategy for three stages of the epidemic: (i) starting phase (blue), (ii) containment after enforcement of interventions (red) and (iii) relaxing of interventions and monitoring for a possible second outbreak (green) (Fig.2). The strategy relies on Bayesian experimental design and can operate when no data are available (as in the start of the epidemic) as well as when data have been accumulated, as in the last two stages of the epidemic. Testing campaigns rely on acquiring randomized samples from a population. The collected data, together with epidemiological models, help determine quantities of interest, such as the basic reproduction number of the disease [14]. By suitably adapting the testing campaign, the data can help reduce the model uncertainty, thus enabling improved estimates regarding the severity of the epidemic.

A testing campaign consists of a set ***s*** of surveys *s*_*i*_ = (*k*_*i*_, *t*_*i*_) which are labeled by *i* = 1, … *M*_*y*_ and performed in locations *k*_*i*_ ∈ 𝒞 and on days *t*_*i*_∈ 𝒯, where 𝒞 and 𝒯 are the set of all available locations and days, respectively. In this paper a survey aims to determine the number of unreported infectious individuals in a particular location on a particular day. In the following we assume limited testing resources, where *N* test-kits are available and each testkit corresponds to testing one person. The goal is to allocate these test-kits in different times and locations so that we maximize the information gain regarding forecasts of the epidemiology model. The locations are the different Swiss cantons, and 𝒞 := {ZH, BE, LU, …} is the set of the strings with canton name abbreviations.

The results of the survey in a canton enable the estimation of a desired quantity of interest, such as the size of the unreported infected population (*I*^*u*^). The number of samples needed to estimate population proportions within a given confidence interval, error tolerance, and probability of proportion is given by Cochran’s formula [9] corrected for a finite population size. Using Cochran’s formula with confidence level 99%, error tolerance 1% and probability of infection 0.1 we find that the samples that would be required to survey the largest Swiss canton (of Zurich) are approximately 5950. All the other cantons need up to 14% less samples with the exception of the smallest canton that needs 27% less samples (figure S7 of the Supplementary Information). Hence we assume the minimum sample size is the same for all cantons. Assuming random sampling of a population with higher probability (up to 0.9) of infection or requiring tighter error bounds, would have implied even more samples according to Cochran’s formula. We note that as of October 2020, 1500 tests per one million people are performed on a daily basis in Switzerland [16]. This amounts to approximately 460 individual tests per canton, which is about an order of magnitude less than what would be required from Cochrans’s formula for an informative random sampling. In turn, by using the proposed OPALITS we can compensate for this lack of test kits with an optimal and systematic process.

We outline the application of the proposed approach to a country with distinct administrative units (cantons in the case of Switzerland) (see figure 1). First, we determine how many cantons will be surveyed, given the number of available test-kits *N*. Then, the sequential optimization of the expected utility function is performed (see Materials and Methods) to identify optimal survey locations (cantons). We then distribute the test-kits to the identified cantons and test a random subset of their population on the suggested day. After collecting the results from all the surveys we update the prior distributions of the model parameters. The collected data leads the maximal information gain in the model parameters. This in turn translates into minimal uncertainty in predictions made with the model for quantities such as the number of unreported infections.

**Figure 1:**
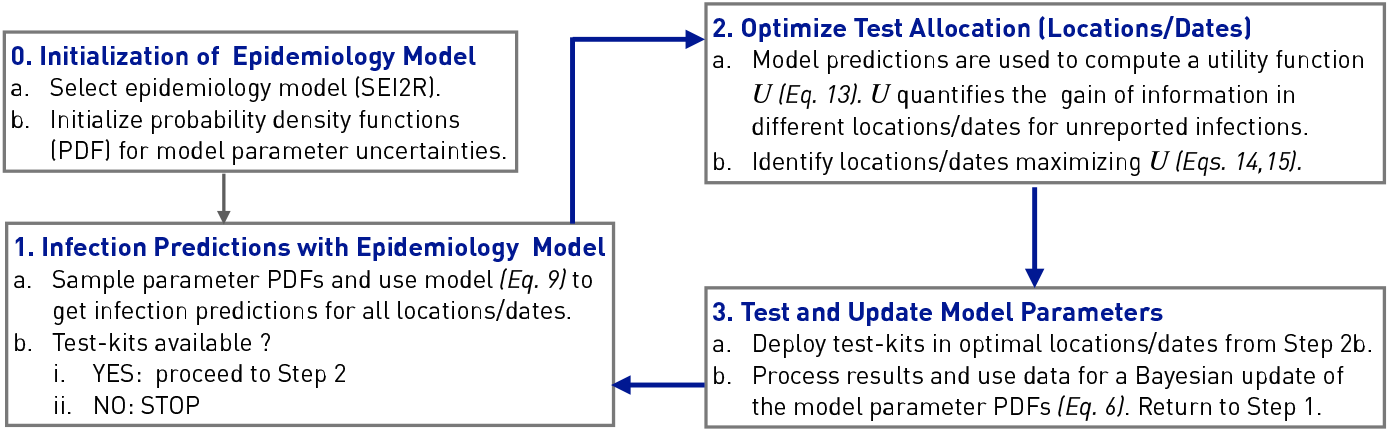
Schematic for the deployment of the Optimal Allocation of Limited Testing Resources (OPALITS) methodology.

**Figure 2:**
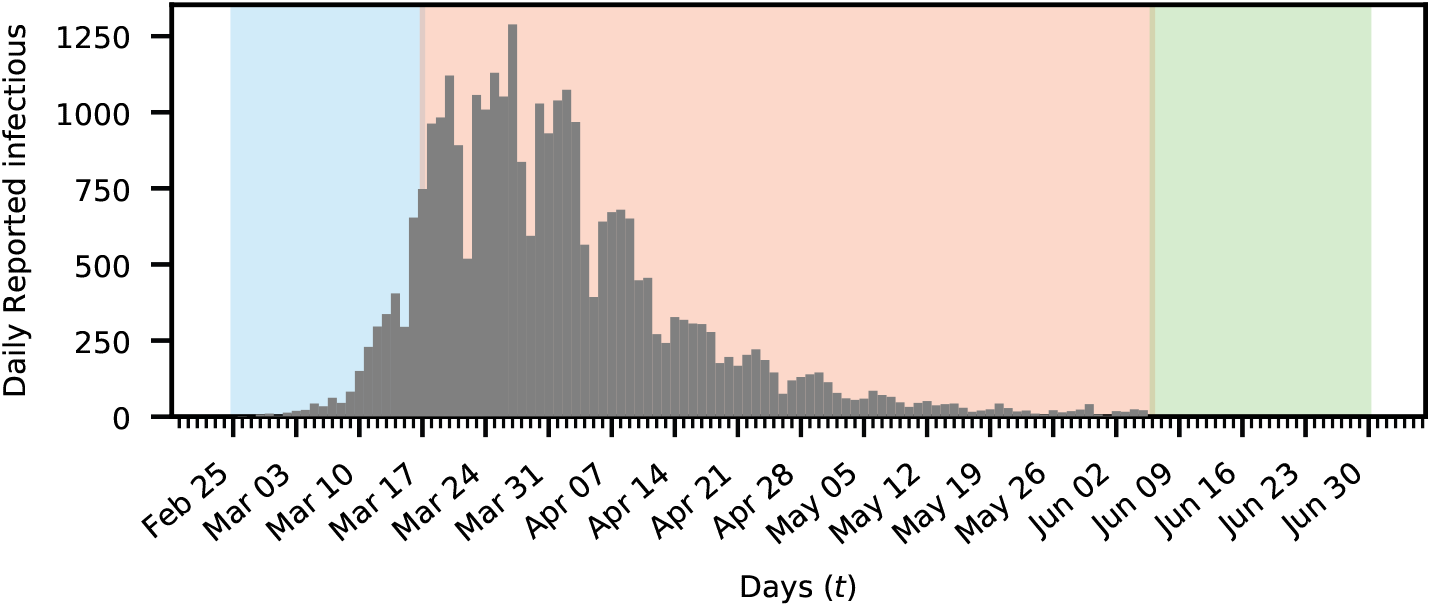
Testing scenarios for the COVID19 outbreak in Switzerland. Daily reported Coronavirus cases in Switzerland are plotted as gray bars. The period before (blue), during (red) and after (green) imposing non-pharmaceutical interventions are marked with color.

The expected information gain of a particular strategy for selecting the survey locations/times ***s*** is quantified by a utility function *Û*(***s***) [15]. The maximum of this function corresponds to an optimal strategy that yields the most information about the quantities of interest. The expected utility function can be understood as a measure of the difference between prior knowledge of the model parameters and the posterior knowledge, after surveys have been conducted in a set of locations and dates. Given such a set, the utility function estimates the expected difference, equivalently the information gain, by taking the expectation over all possible survey results.

The OPALITS relies on forecasts by suitable epidemiological models. In turn, these forecasts rely on prior information and their predictions are further adjusted as data become available in a Bayesian inference framework [17]. The set of Ordinary Differential Equations (ODEs) describing the *SEI*^*r*^*I*^*u*^*R* model [13] are integrated to produce the model output. The uncertainty of the model output and its discrepancy from the available data is quantified through a parametrized error model. The resulting stochastic model and its quantified uncertainties are then used to identify the optimal spatio-temporal allocation of limited test resources.

#### Case 1: Beginning of the epidemic - Optimal testing without data

At the start of an epidemic, there are no data and we assume no other prior information regarding the spread of the pathogen in a country. The initial conditions for the number of unreported infections 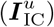 were selected with non-zero values for the cantons of Aargau, Bern, Basel-Landschaft, Basel-Stadt, Fribourg, Geneva, Grisons, St.Gallen, Ticino, Vaud, Valais and Zurich based on their population and their large number of interconnections. Due to the lack of any prior information and relevant data, all the parameters are assumed to follow uniform prior distributions (see table S5, for details).

The first infectious person in Switzerland was reported on February 25^th^ in the canton of Ticino 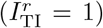 with no initial reported infections in all other cantons. The initial number of exposed individuals is set proportional to the number of unreported infections 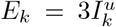 in accordance with the value of *R*_0_ ≈ 3 reported in [18] in the initial stage of the disease. The rest of the population is assumed to be susceptible. The methodology involves parameters of interest (***ϑ*** = (*β, µ, α, Z, D, θ, c*)) and nuisance parameters 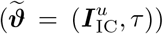) that the testing strategy does not aim to determine (see Materials and Methods section for definitions).

The estimated expected utility functions *Û*(***s***) for up to four surveys in the cantons of Switzerland for a time horizon of 8 days is shown in Figure 3, 𝒯 = {Feb 25, …, Mar 3}. Higher values for expected utility are estimated in cantons with larger population reflecting the larger relative uncertainty for cantons with only few reported cases. This implies that smaller cantons, with lower mobility rates, are less preferred for performing tests since their contribution to the information gain is not significant. This is reflecting the fact that the assumed covariance matrix is shared among cantons (see Materials and Methods). This implies a smaller relative error, when surveying larger cantons with consequently higher number of infections. The Bayesian analysis enables the inference of the particular cantons and days for which a survey should be performed in order to maximize the information gain. Accordingly, the most informative survey should have been made in Zurich on March 2^nd^. The optimal location and time for the second survey is determined to be canton of Vaud on the 27^th^ of February. As expected, the information gained from tests in the canton of Vaud is less than the information gained from the canton of Zurich. The information that would have been gained by surveying the next two selected cantons of Vaud and Basel-Landschaft on March the 3^rd^ and February the 28^th^ respectively, is progressively reduced to a small level that, given the testing costs, does not justify carrying out surveys in more than 4 cantons. The values of the optimal times are listed in table S1 in the Supplementary Information.

**Figure 3:**
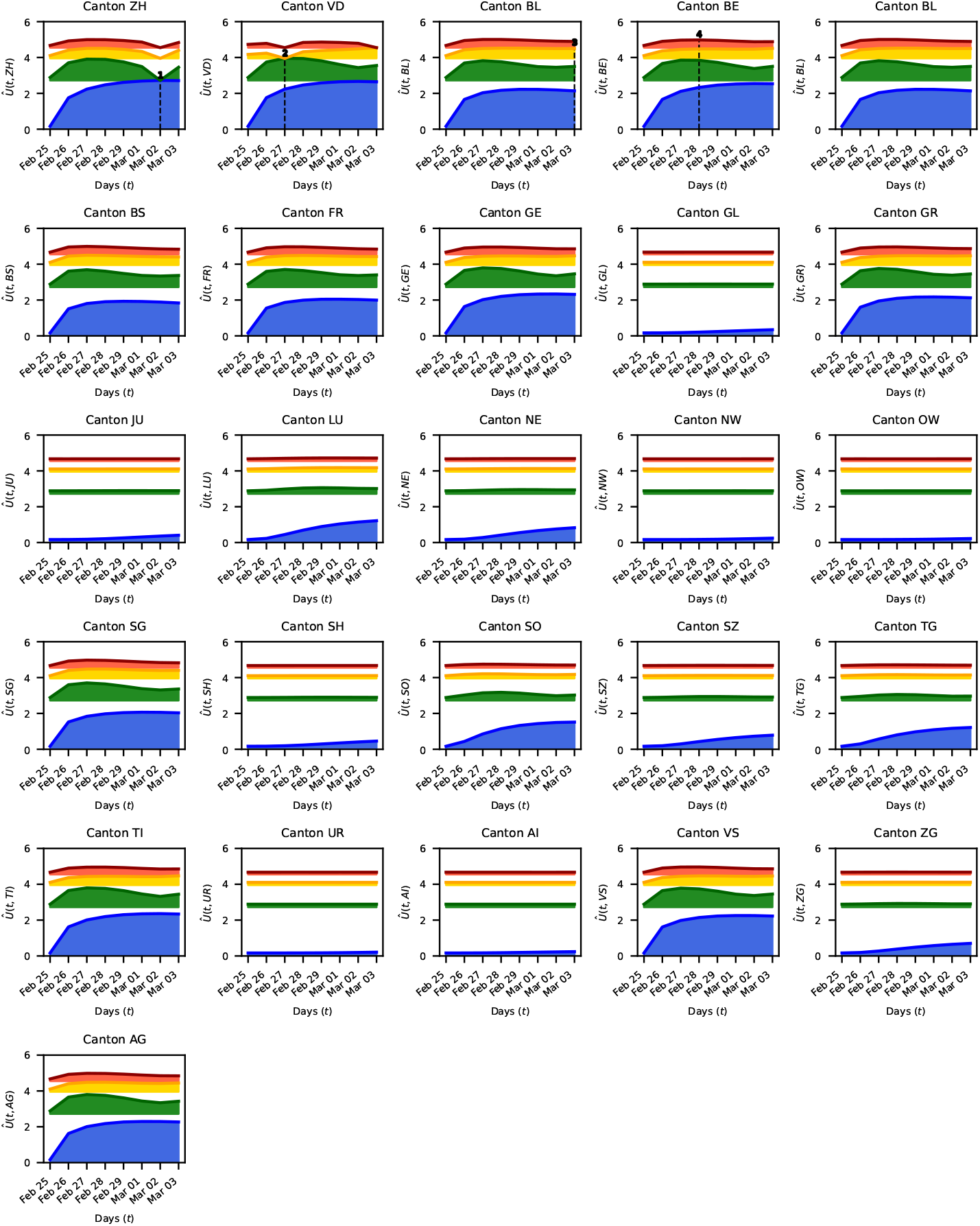
Expected information gain during start of epidemic. The blue curve corresponds to the utility of making one survey. The green curve is the utility when a second survey is added, provided the location and time of the first survey correspond to the maximum of the blue curve (found in the canton of Zurich, on March 2^nd^). Similarly, the yellow and red curves show the utilities for a third and fourth surveys, when the locations and time of the previous surveys are fixed to their optimal values. The fixed dates and location of each survey are plotted with black dashed lines. The shaded areas indicate the difference to the expected information gain of the previous survey, which becomes thinner as additional surveys do not yield a further significant information gain.

The results indicate that the proposed OPALITS methodology selects certain populous and well interconnected cantons at specific times to acquire the most information for estimating the model parameters.

#### Case 2: Exponential spreading and optimal testing strategy during non - pharmaceutical interventions

When the spreading of the coronavirus entered an exponential growth stage, several governments (including the Swiss) decided to take non-pharmaceutical interventions such as requesting social distancing, closing schools and restaurants, or ordering a complete lockdown in order to contain the epidemic. Here, the goal of the OPALITS is to propose surveys that would help to better assess the effectiveness of these interventions.

In this case, probability distributions of model parameters are informed using data from the existing spread of the COVID-19. The daily reported infections in Switzerland [19] from the 25^th^ of February up to the 17^th^ of March 2020 are used to update the distributions, specified in the previous phase, by using Bayesian inference. The marginal posteriors are plotted in figure S1 of the Supplementary Information. The *SEI*^*r*^*I*^*u*^*R* models the non-pharmaceutical interventions with a time-dependent transmission rate *β* and mobility factor *θ*. These parameters are calibrated by the data and provide an estimate on the timing and effectiveness of the interventions[8].

Figure 4 shows the maximum values of the information gain for each survey for 𝒯 = {Mar 17, …, Mar 30}. For cantons with a small population and low connectivity to other cantons a low information gain is found. The opposite can be observed for cantons with large population and strong connections to other cantons. The values for the maximum utility in time for the measurements are listed in Table S2. If only a single canton were to be selected(due to limited availability of test-kits in the country), then a survey in the canton of Vaud carried out on the 30^th^ of March were to be preferred over surveys in either of the cantons of Zurich, Bern or Geneva (blue in figure 4). If two surveys could be afforded, the OPALITS methodology proposes to carry them out in the same canton (Vaud) on the 17^th^ and on the 30^th^ March (blue and green in figure 4). Note that the canton of Zurich, ranked as the next preferred canton for a single survey (blue in figure 4), is not selected by the methodology since part of the information that would be gained from testing is already contained in surveys performed in Vaud. In case more test kits were available, in addition to the two tests in Vaud, the optimal location and time for a third survey would have been the canton of Grisons on the 30^th^ of March (yellow in figure 4). The canton of Zurich is proposed as the fourth location to be surveyed on the 30^th^ of March as well. However, the information gain from the fourth survey in the canton of Zurich is approximately 10% of the total information gained from the surveys carried optimally in the first three cantons.

**Figure 4:**
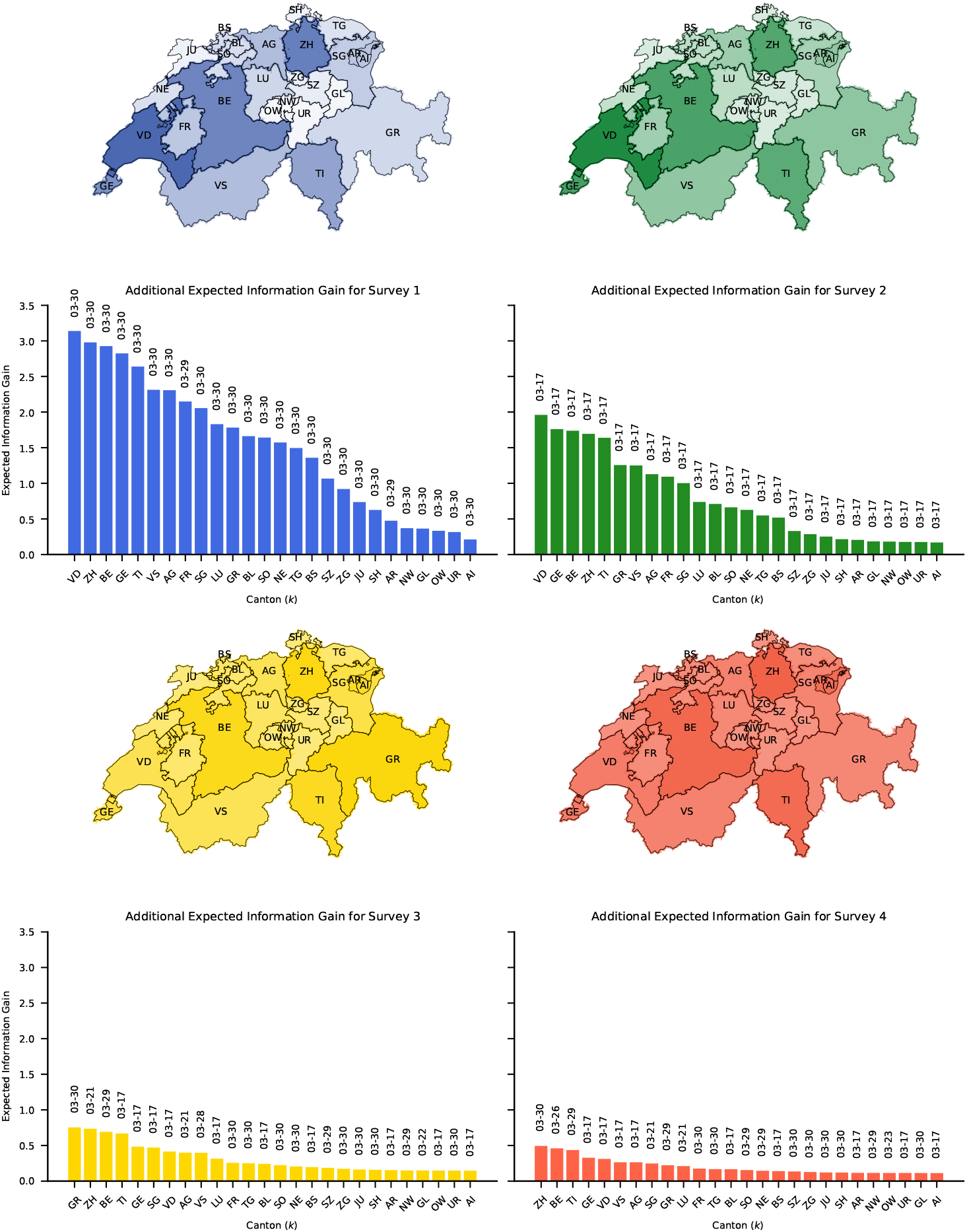
Optimal testing strategy for effect of non-pharmaceutical interventions. The maximum gain of information is plotted on the map of Switzerland using an exponential colormap. Here blue corresponds to taking one survey, green to adding a second, yellow to a third and red to a fourth. Below the map we plot the magnitude of the expected information gain of each survey, along with the optimal measurement dates per canton.

The results suggest that surveys in two locations/times provide significant information regarding assessing the effectiveness of interventions. Further tests on more locations/times did not add substantial information. It is evident that a trade-off between the required information gain and cost of testing are decisive for the number of necessary surveys and respective test kits.

#### Case 3: Optimal monitoring for a second outbreak

After the relaxation of measures that assisted in mitigating the initial spread of the disease, it is critical to monitor the population for a possible second outbreak. The OPALITS methodology supports such monitoring with surveys of the population based on data up to and after the release of the measures.

First, Bayesian inference is performed with data available from February the 25^th^ up to June the 6^th^, to update the uniform priorsthe resulting marginal posteriors are shown in figure S2 of the Supplementary Information. This date is in accordance to the first stage of major release of measures in Switzerland [20]. The effects of interventions are modeled by a parametrized time-dependent transmission rate and mobility factor (see Materials and Methods). The inferred probability distributions of these additional parameters are taken into account as the OPALITS maximizes the information gain. Note that 𝒯 = {Jun 7, …, Jun 14} in this case.

Subsequently, data from February the 25^th^ up to July the 9^th^ are included, repeating the Bayesian inference and estimating the marginal distributions and predictions shown in Figures S3 and S4, 𝒯 = {Jul 10, …, Jul 17}. The results indicate that the relaxation of measures correlates with an increase in the number of reported infections (Figure 5). The information gain for each canton indicates the most informative surveys should be performed a week after performing the inference. The provided information could then assist in estimating the severity of a second outbreak as indicated by the maximum of the utility in time (Tables S3 and S4). Given that tests should be carried out in four locations and times, the methodology promotes optimal surveys for two different times, within a week, in the cantons of Zurich and Vaud. First, surveys should be performed in Zurich, providing high information gain for both considered cases. The next two surveys are to be performed in Zurich and Vaud, with a rank that depends on the considered case, while the fourth test should be performed in Vaud. We find that the information gain from the last test is approximately 10% of the cumulative information gain from the first three surveys. The number of surveys can be then selected according to the available test-kits *N*.

**Figure 5:**
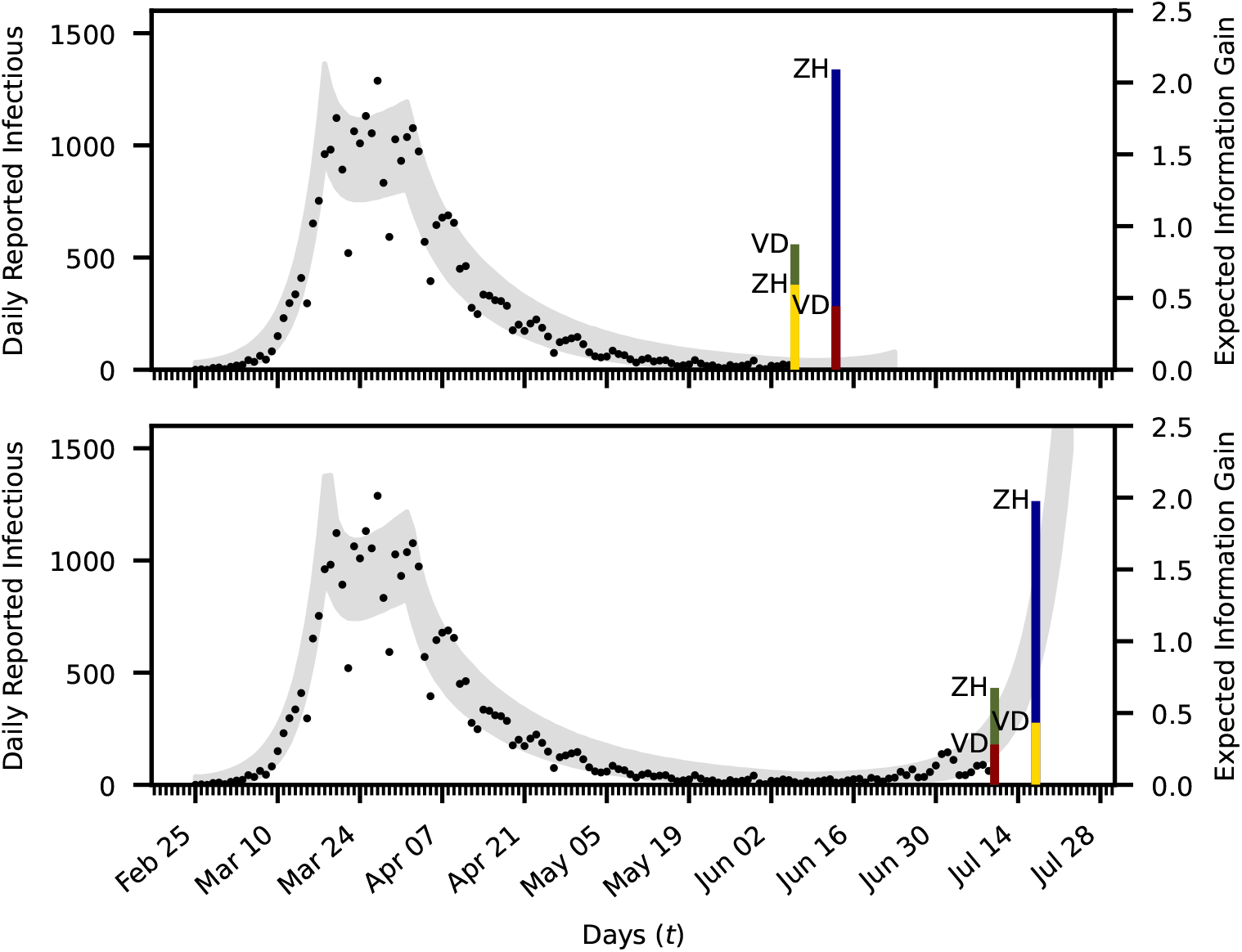
Optimal testing strategy to monitor a second outbreak. Bayesian inference determines the parameters of the first infection wave using the data (black dots) of the daily new reported infections up to the 6^th^ of June (upper plot) and to the 9^th^ of July (lower plot). The 99% confidence intervals are plotted in gray. The proposed testing strategy is plotted with vertical bars at the found optimal days. Here blue indicated the utilities for the first survey. The green bars correspond to the gain in utility when adding a second survey assuming the first was chosen in the optimal location, where the yellow and red correspond to adding a third and fourth survey.

#### Case 4: Effectiveness of Optimal Testing

We demonstrate the importance of following the OPALITS by comparing it with a non-specific testing campaign that is based on heuristics. We first re-examine the situation at the start of an epidemic and assume that the available resources allow for two surveys. Surveys are simulated by evaluating the epidemiological model with the maximum a-posteriori estimate (MPE) of the parameters obtained from the inference in phase II (exponential growth) of the epidemic. We used data of the first 21 days of the infection spread in Switzerland [19] (February 25^th^ to March 17^th^). After evaluating the model, artificial surveys are obtained by adding a stochastic error term.

For the optimal strategy, data are collected by consulting figure 3. Thus, the two surveys are performed in the cantons of Zurich and Vaud, on the 2^nd^ of March and the 27^th^ of February respectively. For a non-specific strategy, the cantons of Ticino and Bern were selected, on the 28^th^ of February. We remark that this is the canton where the first infection was reported and the capital of the country. These artificial data, obtained for the two strategies, are added to the real data of the daily reported cases from the first 8 days after the outbreak in Ticino. For the expanded data-set 𝒟 the posterior distributions 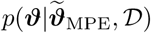 are found by sampling the model parameters using nested sampling [14].

The resulting one- and two-dimensional marginalized posterior distributions for both strategies are shown in figure 6. We note that the dispersion coefficient *r* (defined in the Materials and Methods) in the error model for the real data (the reported infections) and the correlation parameter are almost the same for both strategies. However the model parameters show significant differences even when only two new data-points are added to a set of 208 data-points. The posterior distributions of the parameters of interest are propagated through the epidemiology model to provide the uncertainties in the number of unreported infectious individuals. In figure 7 the model output for the total number of unreported infections is plotted together with a 99% confidence interval along with the true value of the unreported cases obtained by using the selected parameters. The predictions from the OPALITS have a much higher certainty with a confidence interval that is up to four times narrower than the one from a non-specific strategy. The same figure also shows the relative histogram plots for the effective reproduction number, which for the employed model is given from *R*_*t*_ = *βDα* + *βDµ*(1 − *α*) [13]. Not only is the histogram more peaked, when data is optimally collected, but also the mean value of the two histograms is different. When data is optimally collected, the found mean value for the effective reproduction number is 2.1, whereas when the non-specific strategy is followed the average value is 3.2. A mean value of 3.2 could lead to more strict non-pharmaceutical interventions, which might prove unnecessary and harmful for the economy.

**Figure 6:**
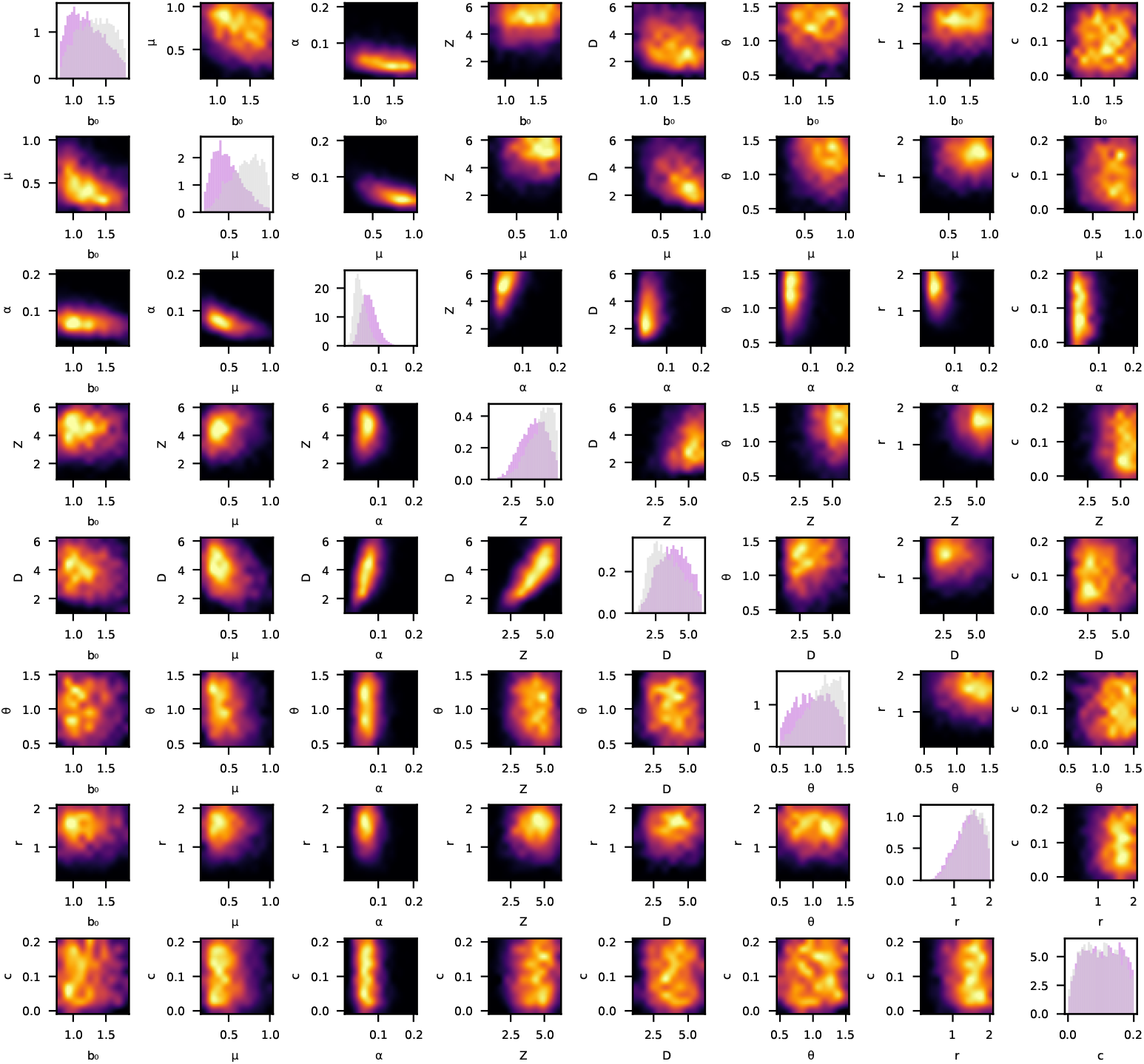
Marginal posterior distributions for two strategies. The diagonal shows the histogram for the marginal distribution for every parameter. Purple indicates posterior for the survey following the optimal testing strategy, gray the one for the non-specific strategy. The lower half and upper half show the samples of the joint distribution of two parameters for the optimal and the non-specific strategy respectively. Here black indicates low density and yellow high density.

**Figure 7:**
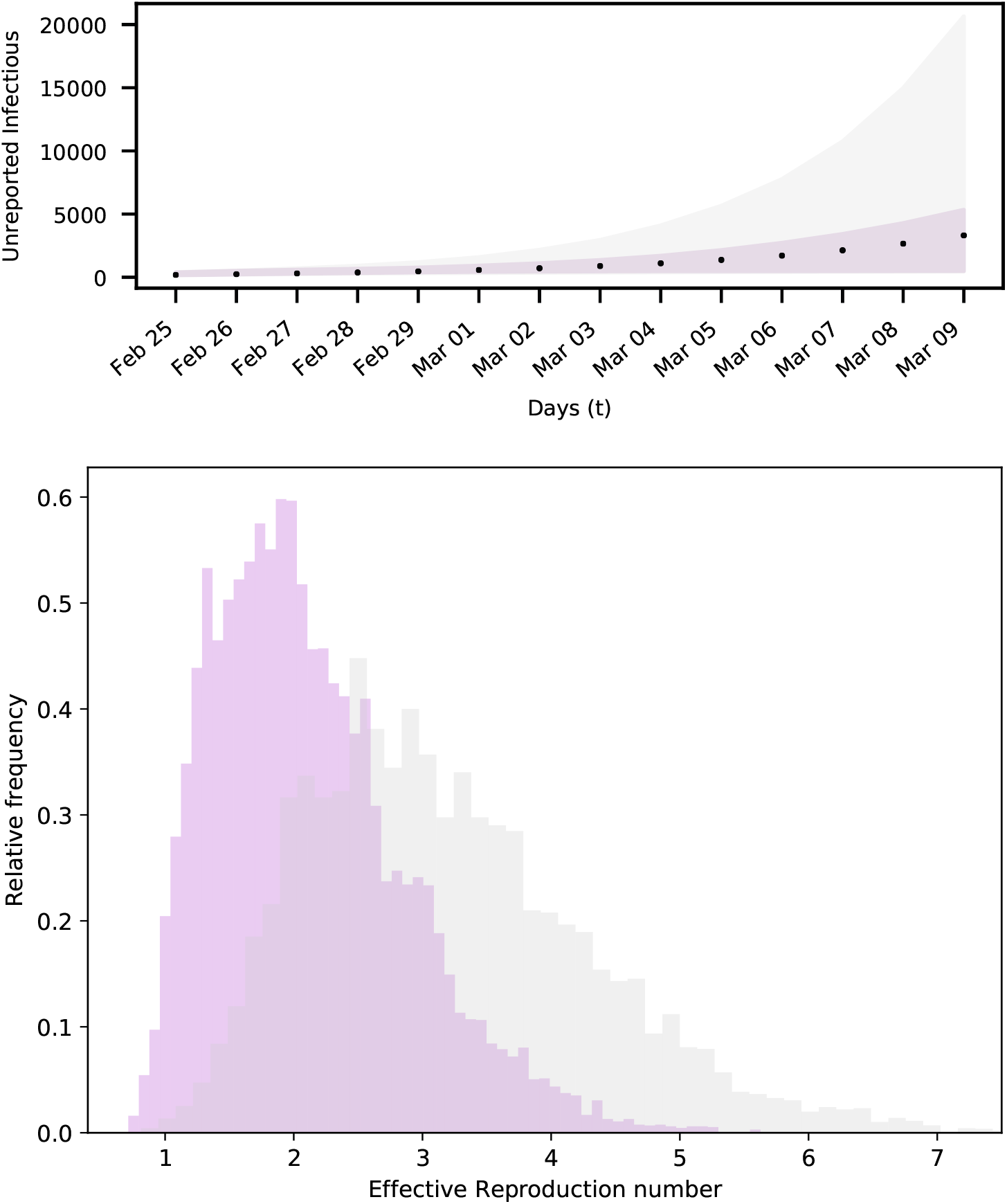
Prediction uncertainty for different testing strategies. Up: The black dots show the actual unreported infectious for an artificial spread in Switzerland. The error bounds show the 99% confidence intervals of the model output for samples of the parameters with data obtained by optimal (purple) and non-specific testing (gray). Down: Relative frequency histograms for effective reproduction number, predicted with data obtained by optimal (purple) and non-specific testing (gray).

Further comparisons, demonstrating the value of the OPALITS, include model predictions with higher certainty, as indicated by confidence intervals that are narrower than the ones obtained from from a non-specific strategy (figures S5 and S6, see Supplementary Information). Narrower uncertainty bounds provide higher confidence for decisions related to possible interventions to contain the epidemic.

## 3. Discussion

We introduce a systematic approach to identify optimal times and locations for epidemiological surveys to quantify infectious individuals in a country’s population during the COVID-19 epidemic. The proposed OPALITS methodology exploits prior information and available data to maximize the expected information gain in quantities of interest and to minimize uncertainties in the forecasts of epidemiological models.

The present study addresses the need for an accurate assessment of COVID-19 infections [21] and it is shown to be far more accurate than the currently applied random testing. The proposed methodology is, to the best of our knowledge, the first method to propose an optimal spatio-temporal allocation of limited test-kit resources. A first study for the estimate of unobserved COVID-19 infections [5] in the USA indicated that early testing would have decreased the surveillance gap during a critical phase of the epidemic. More recently a number of studies have emerged that address the optimal allocation of resources. The “Test and Contain” process suggested in [7] addresses an idealized population of 10’000 and solves an allocation problem using predictions of the SIR model. They assume an isolation of the positively identified individuals and showed that just one test a day can reduce the peak of infected individuals by 27%. This study is similar to ours in casting the test allocation problem in an optimization framework using linear programming in contrast to information maximization that we propose. However, their approach is not data informed and does not address a realistic country scenario. Another study [22] focused on test-kit allocation in the Philippines. They use a statistical approach and non-linear programming to determine the optimal percentage allocation of COVID-19 test-kits among accredited testing centers in the Philippines aiming for an equitable chance to all infected individuals to be tested. Their goal of optimal percentage allocation differs from ours that is optimal space and time allocation of test-kits.

The proposed method is demonstrated by focusing on the outbreak of the epidemic in Switzerland. We compare OPALITS with random testing and demonstrate its advantages in producing forecasts with far reduced uncertainties. We note that the existing testing capacity of 1500 tests per million people in Switzerland can be better allocated than the ongoing random testing. Moreover we show that the present methodology will be of particular importance to countries with testing capacity that is far lower than that of Switzerland [16].

The methodology relies on Bayesian experimental design using prior information and available data of reported infections along with forecasts from the *SEI*^*r*^*I*^*u*^*R* model. We compute the optimal testing strategy for three phases of the epidemic. At the onset of the epidemic the method identifies the most crucial dates and locations for randomized tests in the country’s population. The deployment of OPALITS at this phase would have allowed authorities to perform randomized testing in a period of high uncertainty, well in advance of the disease outbreak. Moreover, the presented approach is applicable to any newly arising epidemic and can be used to identify important surveying locations and a general protocol of action, whenever an unknown disease starts to spread. In the case of COVID-19, such course of action would limit early inaccurate estimates of metrics such as the virus mortality rate, estimated around 3% in early March 2020 by the World Health Organization [23] and currently believed to be lower than 1%, [24].

During the period of non-pharmaceutical interventions the proposed strategy would help quantify their effectiveness assisting decision making for further interventions or retraction of measures that may be harmful to the economy. In this study, available data for the daily reported infections prior to any interventions, combined with the proposed methodology, indicated that conducting two surveys after measures are imposed is sufficient. This can help to identify the new virus dynamics quickly and adjust interventions accordingly. Similarly, the OPALITS can assist monitoring for a recurrence of the disease after preventive measures have been relaxed and help guide further planing of interventions. Since massive testing for a new disease might not be a possibility during its first outbreak and cheap individual tests might become available only later, applying the proposed methodology at this point provides a useful guideline on how to use the individual tests to conduct large-scale surveys. For instance, in Switzerland it was not before mid-April 2020 that rapid COVID-19 tests were released on the market [25]. Collecting data for the reported cases before that and using it to inform the proposed approach to find an OPALITS (after cheap individual tests become available) that will be applied during a possible lock-down would be the suggested course of action in this case.

There are a number of issues that the model should be able to accommodate in the future. These include accounting for virological test sensitivity, delays in the reporting of the test results and bias on the estimate of the unreported infected individuals (Cochran’s formula). Further developments may include models that account for different transmission dynamics in cantons while the classical Bayesian inference methods may be replaced with Hierarchical Bayesian Method to account for heterogeneous data.

We remark that the proposed OPALITS does not depend on a particular type of data/model or to the country of Switzerland. The open source code is modular, scalable and readily adaptable to different scenarios for the epidemic and countries around the world. We believe that the present work can be a valuable tool for decision makers to allocate resources efficiently for testing the population, providing a reliable quantification of the spread of the disease and designing effective interventions.

## Data Availability

The open source software is available on Github.

https://github.com/cselab/optimal-testing

## Acknowledgments

We acknowledge discussions with Fabian Wermelinger, Lucas Amoudruz, Martin Boden (ETHZ). Sergio Martin (ETHZ) provided technical assistance with the software.

## Authors contributions

Conceptualization: C.P., P.Ko.; Data curation: M.C., P.W.; Formal Analysis: M.C., P.W., G.A., D.W., C.P.; Funding acquisition: P.Ko.; Investigation: M.C., P.W., C.P., G.A., P.Ko. Methodology: P.W., M.C., C.P., G.A., P.Ko.; Project administration: P.Ko.; Resources: P.Ko.; Software: P.W., M.C., D.W., I.K., P.Ka.; Supervision: C.P., P.Ko., G.A.; Validation: M.C., P.W.; Visualization: P.W., M.C., P.Ka; Writing – original draft: P.W., M.C., C.P., P.Ko.; Writing – review & editing: P.W., M.C., C.P., P.Ko., G.A., D.W., I.K., P.Ka.

## Data and materials availability

All data is available in the manuscript or the Supplementary Materials. The code is available under https://github.com/cselab/optimal-testing.

## Supplementary Information

### 1. Materials and Methods

The optimal time (day) and location (canton) for surveying a population to detect infectious individuals is determined via Bayesian optimal experimental design [1]. This optimal testing allocation (OPALITS) relies on combining Bayesian inference and utility theory with forecasting models of the epidemic. We remark that the OPALITS does not depend on a particular epidemiological model or type of data. The methodology is applicable at all stages of the epidemic (inception to re-occurrence). It can operate without data at the early stages of the pandemic and takes advantage of data available at later stages of the pandemic. The methodology is rendered computationally efficient using a sequential optimization algorithm [2].

#### Bayesian Inference from randomized testing

We consider a testing campaign including a set (*s*) of surveys *s*_*i*_ = (*k*_*i*_, *t*_*i*_), *i* = 1, … *M*_*y*_ performed in location *k*_*i*_∈ 𝒞 and on day *t*_*i*_∈ 𝒯. These surveys measure a quantity of interest (QoI), that is denoted by ***y***(*s*) = (*y*_1_, …, *y*_*My*_). Here, *y*_*i*_ is the number of unreported infectious individuals, measured through survey *s*_*i*_. The QoI can be predicted by a model 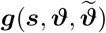 (here the *SEI*^*r*^*I*^*u*^*R* epidemiological model) that depends on parameters of interest ***ϑ*** ∈ ℝ^*N*^ and nuisance parameters 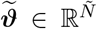. The distinction between model and nuisance parameters is discussed in later sections. We note that both sets of parameters are uncertain and the proposed method aims to reduce the uncertainty only in the parameters of interest.

A stochastic error term *ε*(*s*) links the model prediction with the QoI

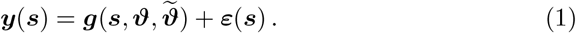

The error *ε*(*s*) is assumed to follow a zero-mean multivariate normal distribution 𝒩 (0, Σ) with covariance matrix 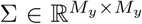. The elements of the covariance matrix (Σ_*s,s*_′) correspond to surveys taken at *s* = (*k, t*) and *s*′ = (*k*′, *t*′) and are given by

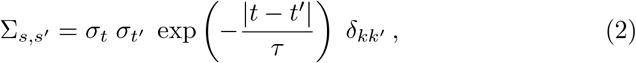

where *δ*_*kk*′_ is the Kronecker delta, which is 1 for *k* = *k*′ and 0 otherwise. The correlation time *τ* ∈ [0.5, 3.5] is considered a nuisance parameter. These assumptions about the covariance imply that surveys in different locations are not correlated, while those in the same location have an exponentially decaying temporal correlation. The latter avoids clustering of surveys in small time intervals [3]. The factor *σ*_*t*_ ∈ ℝ is assumed proportional to the expectation of the QoI, taken over all possible survey locations and over the range of model and nuisance parameters

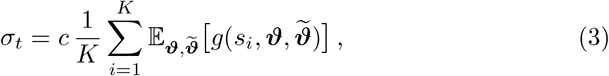

where *s*_*i*_ = (*i, t*). The parameter *c* ∈ [0, 0.25] is considered a model parameter. The expectation 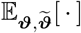 is taken with respect to all parameters ***ϑ*** and 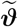 that follow the prior probability distribution with density 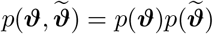.

Under these assumptions, the conditional probability of ***y*** on 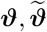 and *s* is given by

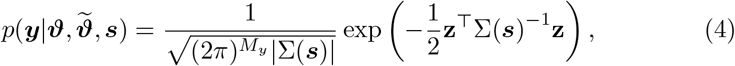

where |Σ(*s*)| is the determinant of the covariance matrix and 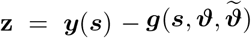.

In the present study, the QoI measured by a survey is the number of unreported infectious individuals in a particular canton on a particular date. This implicitly assumes that there no restrictions on when the survey can be conducted and that there are no observational delays, which means the the QoI is instantaneously obtained. Both assumptions are not restrictive however. Restrictions on the possible survey dates can be accounted for by simply excluding those dates from the dates on which the utility function is evaluated. Also, a delay of one day (meaning that two days are needed to survey a canton *k*, starting from day *t*) would mean that 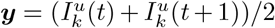 is measured. In other words, when there is a delay the measured quantity can still be mapped to a model quantity, which allows us to perform Bayesian inference. There are several types of measurements (Rapid testing [4], PCR [5], Schwabs [6]) being proposed for testing asymptomatic individuals. We emphasize that our methodology is compatible with any of these types. Data related issues such as uncertainties, test sensitivities and delays in processing can be accommodated in the Bayesian inference framework and in the input to the SEIR model.

#### Expected Information Gain

The most informative surveys ***y*** provide the least uncertainty in the estimates of the model parameters ***ϑ***. Starting with a user-postulated prior distribution *p*(***ϑ***), Bayesian learning is used to update the uncertainties in the model parameters leading to a posterior distribution 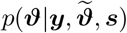, based on the information contained in the test data ***y***. The Kullback–Leibler (KL) divergence between the posterior 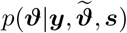 and the prior distributions *p*(***ϑ***) of the model parameters measures the distance between the two distributions. Informative data produce posterior distributions that differ from the prior; greater differences lead to higher information gain. Therefore, the most informative data ***y*** correspond to the testing strategy (measurement locations and times) with the highest information gain [7, 8].

The OPALITS is identified by maximizing a utility function [1]. One choice is the KL divergence 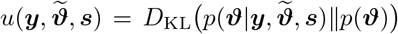 quantifying the information gain from the data [1]. However, since data are not available in the experimental design phase, the utility function is selected here to be the expected KL divergence 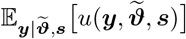 over all data generated by the model prediction error equation 1. Also, to account for the uncertainty in nuisance parameters 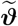, encoded in the prior distribution 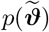, the expectation is also taken with respect to 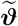, which results in the utility function [1]

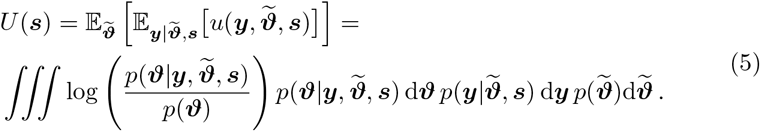

By using Bayes’ theorem

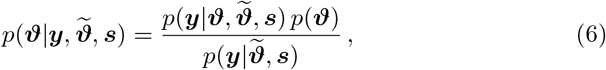

the utility function can be simplified to

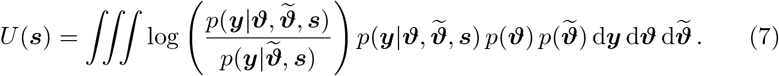

Note that the expected utility only depends on the locations and times of the measurements via *s*. The term 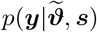 is the model evidence given by

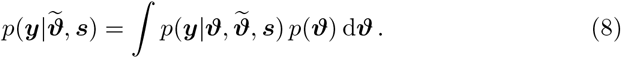

The choice of the prior distribution *p*(***ϑ***) for the parameters allows to incorporate prior knowledge from epidemiology. If no information is available from data, a case encountered in the beginning of the infection, a uniform prior distribution can be assumed. Table S5 summarizes our choice of prior distributions for all the involved uncertain quantities. If data ***d*** of the daily number of reported infectious individuals is available, Bayesian inference can be used to inform the prior distribution, as described later on. In this case, the prior *p*(***ϑ***) in equation 7 is replaced by the distribution *p*(***ϑ***|***d***) informed from the data ***d***.

In the present work, the assumed nuisance parameters are the correlation time *τ* and the initial condition of the unreported infections in the cantons of Aargau, Bern, Basel-Landschaft, Basel-Stadt, Fribourg, Geneva, Grisons, St.Gallen, Ticino, Vaud, Valais and Zurich

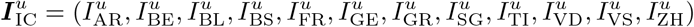

with prior distributions 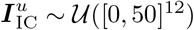 and *τ* ∼ 𝒰([0.5, 3.5]).

#### Epidemiological Model

Here we employ the *SEI*^*r*^*I*^*u*^*R* epidemiological model [9] to forecast the dynamics of the coronavirus outbreak in Switzerland

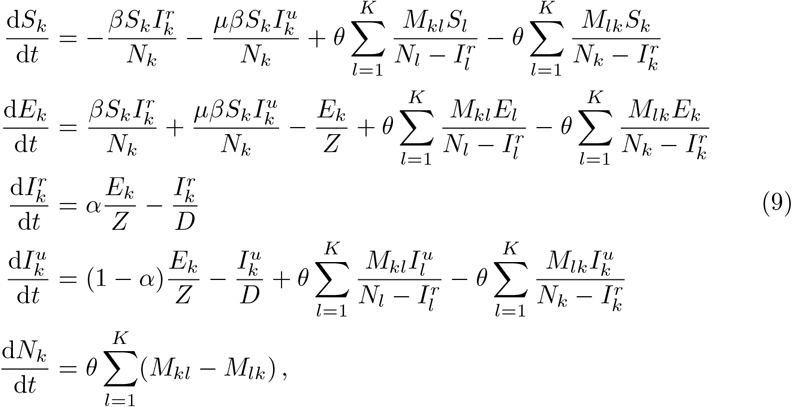

where 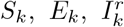 and 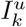 denote the number of individuals in canton *k* = {1, …, *K*} that are susceptible, exposed, reported infectious and unreported infectious, respectively. We denote by *K* the number of cantons (26 in Switzerland), by *N*_*k*_ the total population of the canton *k*, while the population mobility between cantons *k* and *l* is denoted by *M*_*kl*_ with values obtained from the Swiss Federal Statistical Office [10]. The model parameters are the transmission rate (*β*), the relative transmission rate between reported and unreported infectious individuals (*µ*), the virus latency period (*Z*), the infectious period (*D*), the reporting rate (*α*) and the mobility factor (*θ*).

We employ different time-dependent expressions for the transmission rate and the mobility factor for each stage of the epidemic. Constants are chosen for the start of an epidemic while in the cases of monitoring of interventions, the following expressions are used:

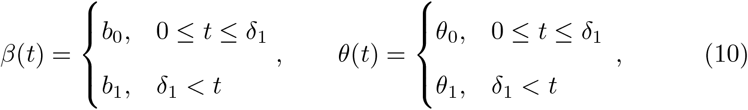

where *b*_0_, *b*_1_, *θ*_0_ and *θ*_1_ are the transmission rates and mobility factors before and after the intervention. Time *t* = 0 corresponds to the 25^th^ of February 2020, and *δ*_1_ = 21 to the 17^th^ of March 2020, when the lockdown was announced in Switzerland [11]. Finally, for the third case (monitoring of a second outbreak) we assume that

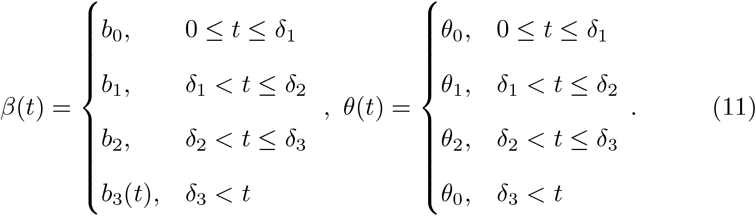

As in equation 10, *b*_0_ is the transmission rate before the intervention while *b*_1_ = *c*_1_ *b*_0_ and *b*_2_ = *c*_2_ *b*_0_ with *c*_1_, *c*_2_ ∈ [0, 1] are the transmission rates after the two interventions. Similarly, *θ*_0_ is the mobility factor before any interventions took place, while *θ*_1_ = *c*_3_ *θ*_0_ and *θ*_2_ = *c*_4_ *θ*_0_ with *c*_3_, *c*_4_ ∈ [0, 1] are the mobility factors after the two interventions. Moreover, *δ*_1_ and *δ*_2_ correspond to the days of the interventions. The day when the measures are loosened is denoted by *δ*_3_. After that day, the transmission rate is gradually increasing

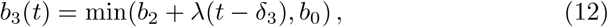

with *λ* ∈ [0, 0.03], while the mobility factor regains its initial value of *θ*_0_.

#### Estimation of the Expected Information Gain

The calculation of the expected utility from equation 7 is performed with Monte-Carlo integration. Samples from the prior distribution are denoted by ***ϑ***^(*i*)^ ∼ *p*(***ϑ***) and by 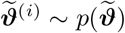, while samples on the measurement space are denoted by 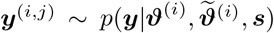, where *i* ∈ {1, …, *N*_*ϑ*_} and *j* ∈ {1, …, *N*_*y*_}. With these samples, an estimate of the expected utility is computed as

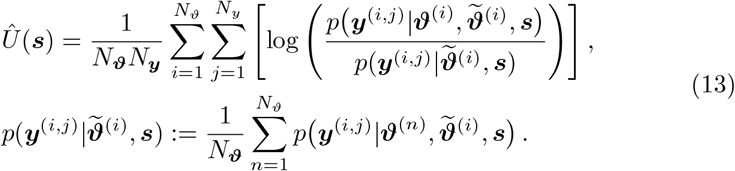

In our implementation the samples ***ϑ***^(*i*)^ and 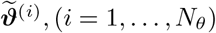, remain the same for different values of *s*. Thus, the model evaluations 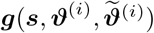 are only carried out once and are stored and used in the iteration process involved in the optimization. This allows to separate the computational cost of the model evaluation from the cost of computing the utility, which scales as 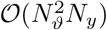.

#### Optimal Location and Time of Testing

We define the optimal survey times and locations as

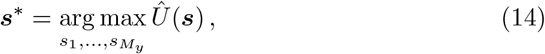

where 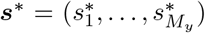 with 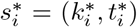 denote the locations 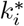 and times 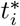 for the optimal surveys with *i* ∈ {1, …, *M*_*y*_}. For a grid search, the associated computational cost is 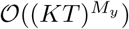 and thus grows exponentially with the number of surveys. This curse of dimensionality is avoided by using a sequential optimization method [2] to approximate the global optimum by iteratively solving

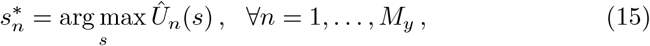

where *s* = (*k, t*) is the location and time to be estimated sequentially starting with *n* = 1 and

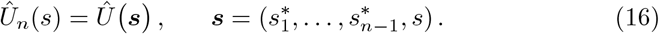

Following this, we define the expected information gain for survey *n* as

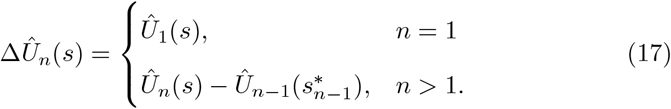

#### Quantification of Uncertainty

A data informed prior *p*(***ϑ***|***d***) of the model parameters ***ϑ*** can be computed from available data 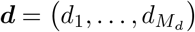, collected at *M*_*d*_ locations and days. Here, available data ***d*** refer to the daily number of reported infectious individuals and they are contrasted from the data ***y*** of the number of unreported infectious individuals. The latter are obtained from testing strategies at selected populations using optimal experimental design. The data is mapped via a distinct model output 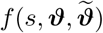 through the following error model

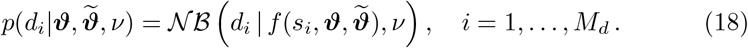

where 𝒩 ℬ is the negative binomial distribution with mean *f* and dispersion *ν*. Also, *s*_*i*_ = (*k*_*i*_, *t*_*i*_) is the location and time the data *d*_*i*_ was collected. The choice of a different error model, compared to equation 1, is based on the assumption that the data are independent and identically distributed. Such an assumption would not be acceptable in the measurement model in equation 1, as it may result in uncorrelated measurements that can become clustered in small time intervals [3].

The data 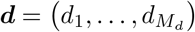 are the daily number of reported infections per canton in Switzerland [12] which corresponds to the following model quantity

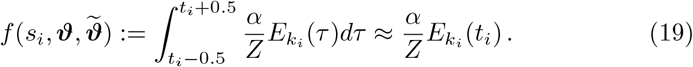

The posterior distribution that will be used subsequently as a data informed prior is obtained using Bayes’ theorem

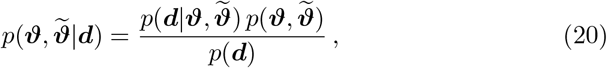

and is sampled with a nested sampling algorithm [13]. Note the difference to equation 6 and the optimal testing methodology, where we are interested to reduce the uncertainty in 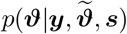, which excludes the nuisance parameters 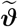. For the dispersion parameter in equation 18, it is assumed that 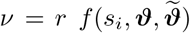. The coefficient *r* is unknown and included in the parameter set, where *r* ∼ 𝒰([0, 2]).

**Figure 1:**
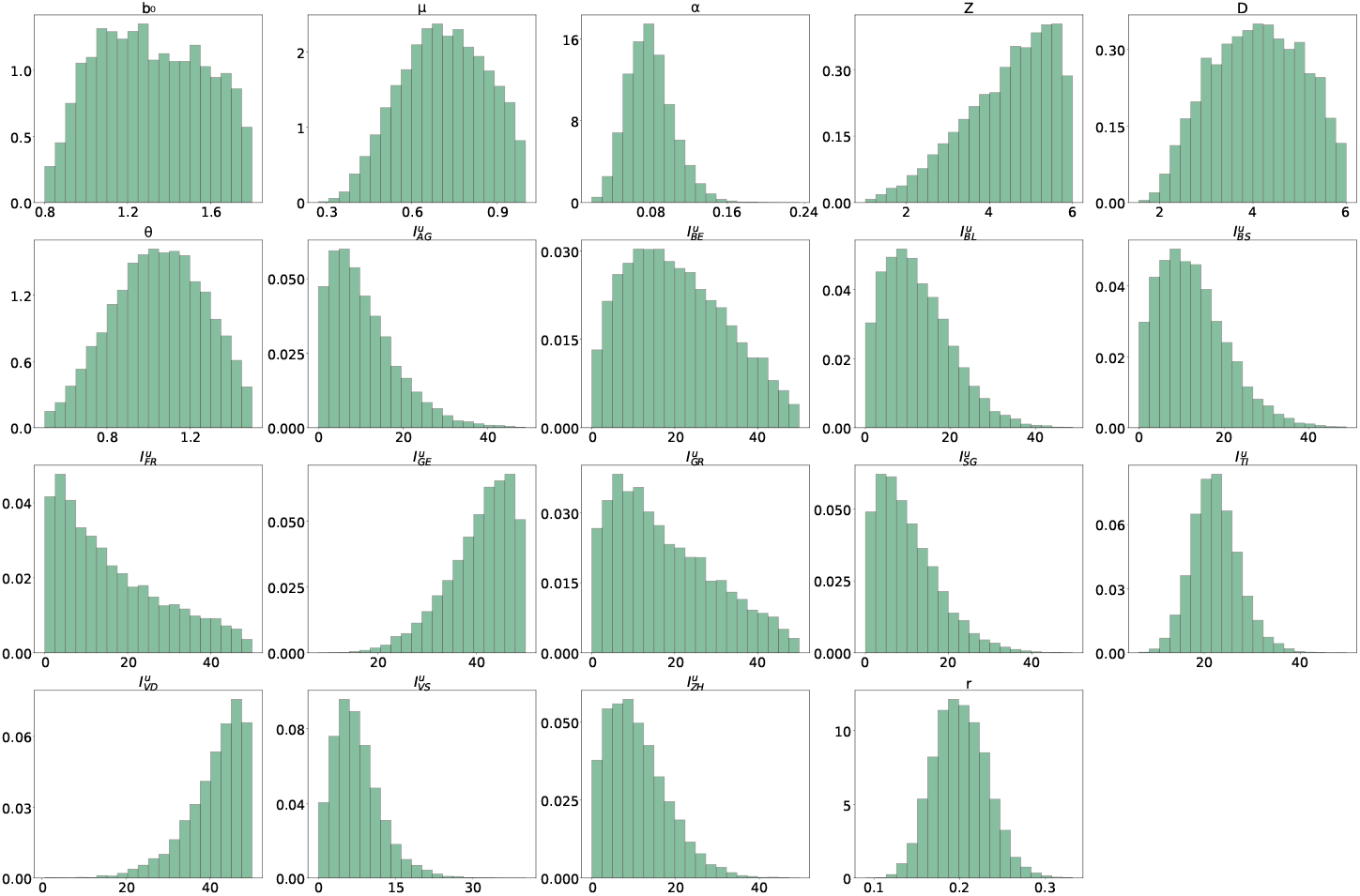
Marginal posterior distributions with data up to 17^th^ of March 2020. The used data correspond to the daily reported infectious persons in the cantons of Switzerland. The marginals with a canton label XY correspond to the initial condition 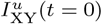 for the unreported cases in that canton.

The three inferences performed are summarized in table S5, which shows the involved model parameters in each case. The histograms for the found samples are shown in figures S1, S2, and S3.

We remark that, using the present methodology, the inferred date for the beginning of the intervention is *δ*_1_ = 22.5, which is the 18^th^ of March 2020, corresponding well with the 17^th^ of March 2020 on which the lockdown was introduced in Switzerland [11]. Moreover, we infer a significant reduction in the mobility factor, which indicates that traffic between cantons was also minimized. For the inference III we plot the fit using the inferred parameters in figure S4. The daily reported cases per canton are shown, together with the data used for the inference.

**Figure 2:**
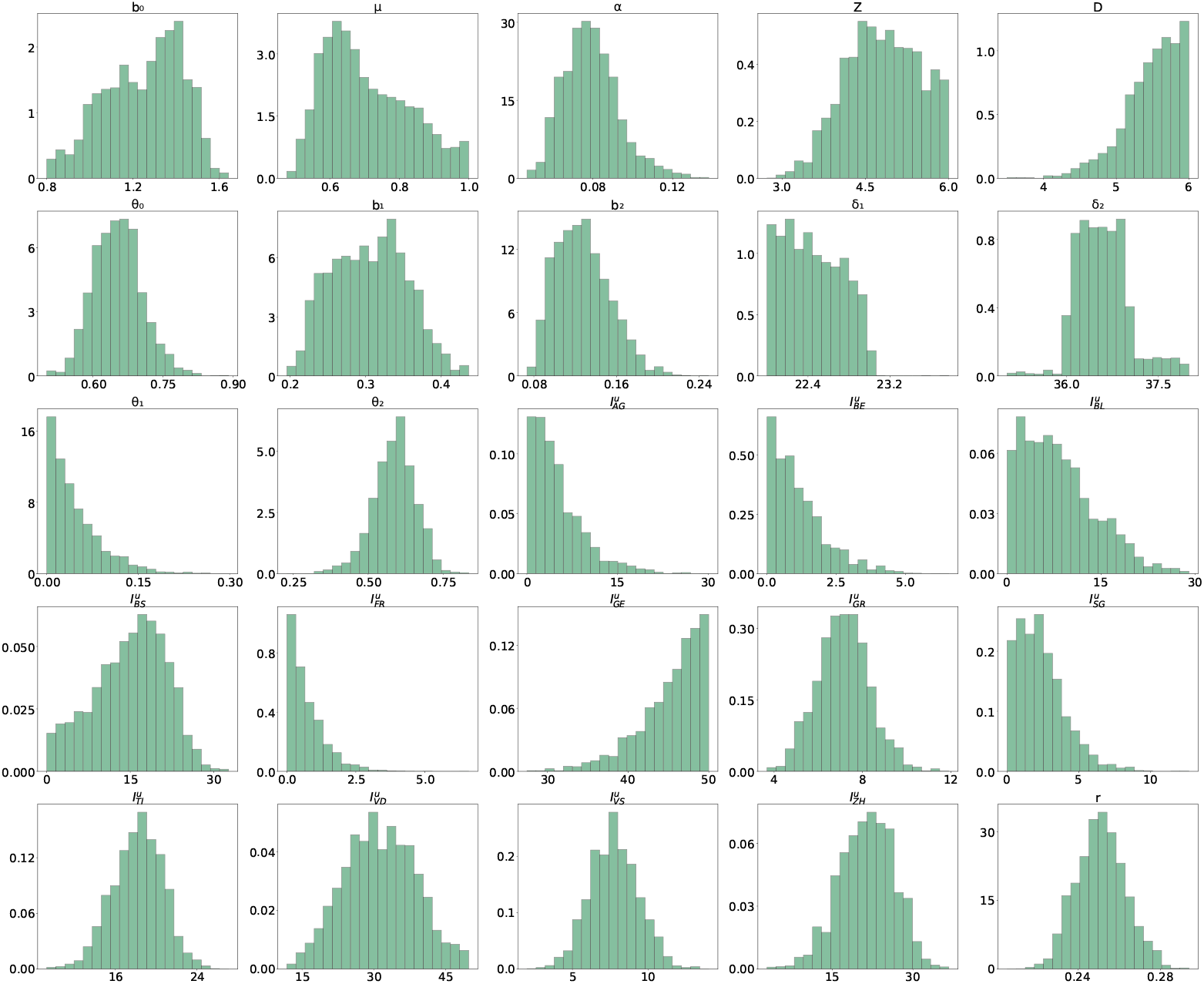
Marginal posterior distributions with data up to 6^th^ of June 2020. The used data correspond to the daily reported infectious persons in the cantons of Switzerland. The marginals with a canton label XY correspond to the initial condition 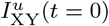 for the unreported cases in that canton.

**Figure 3:**
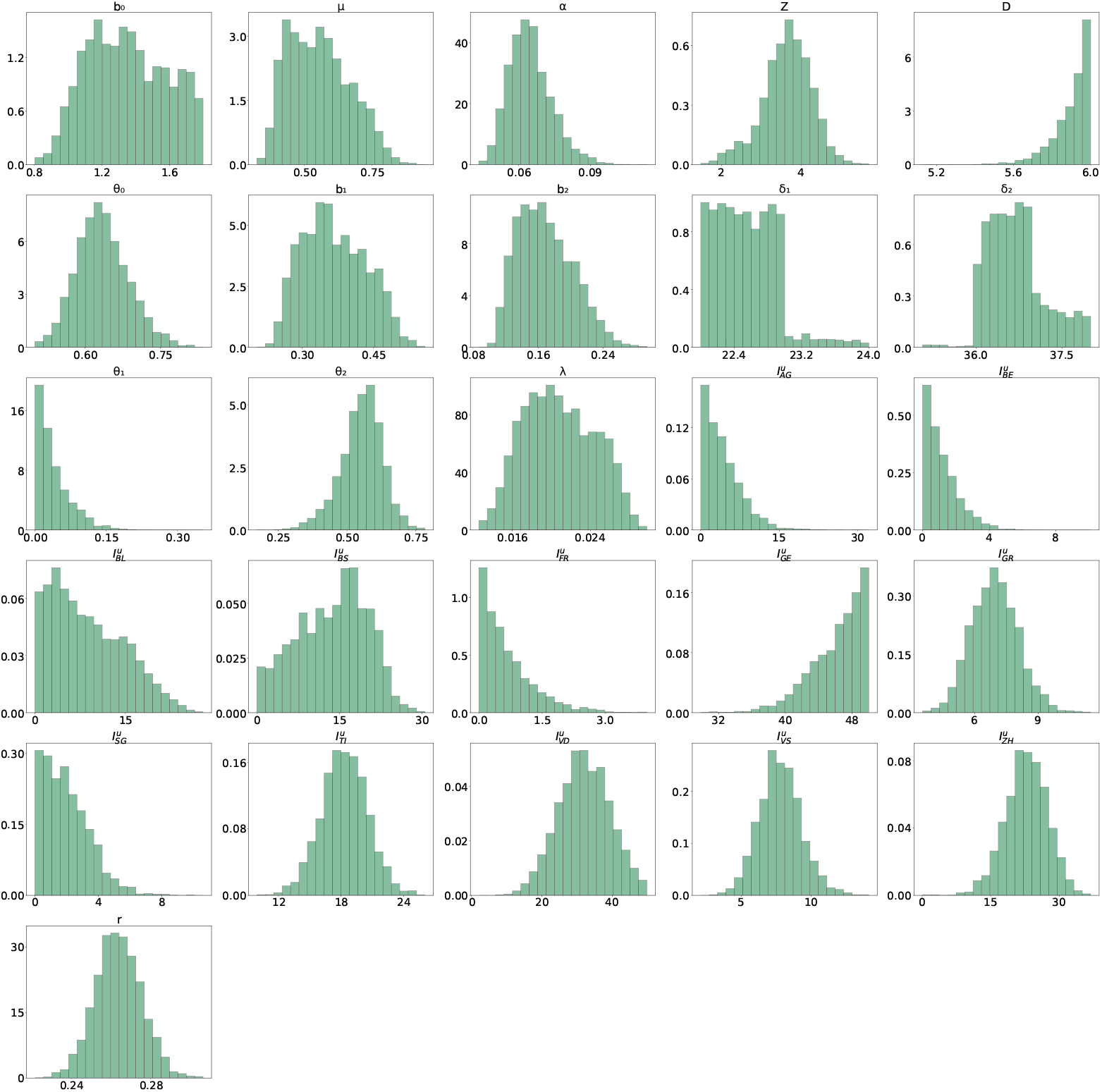
Marginal posterior distributions with data up to 9^th^ of July 2020. The used data correspond to the daily reported infectious persons in the cantons of Switzerland. The marginals with a canton label XY correspond to the initial condition 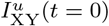 for the unreported cases in that canton.

**Figure 4:**
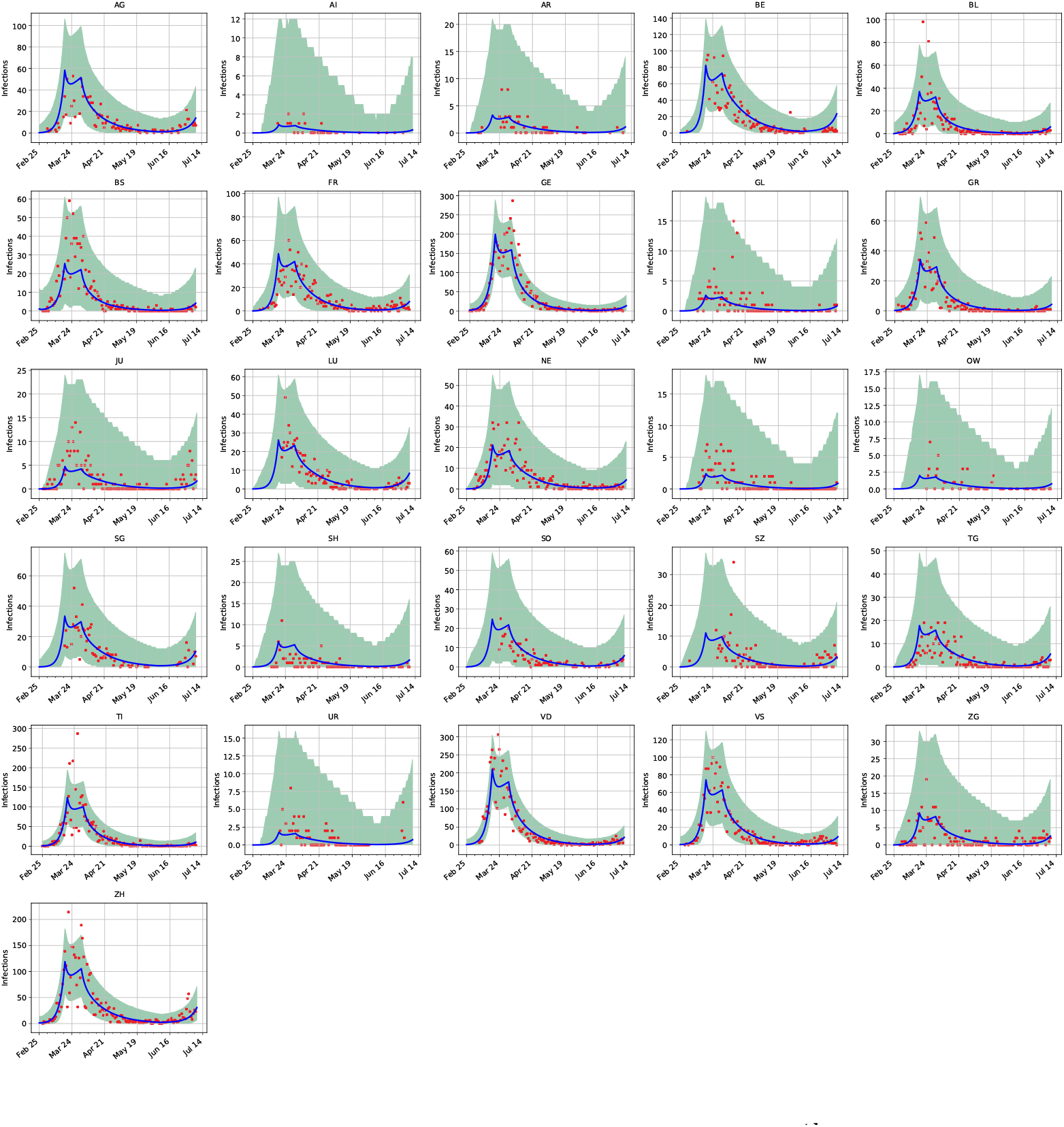
Maximum a-posteriori prediction with data up to 9^th^ of July 2020. The red points correspond to the daily reported cases per cantons and the blue curve shows the maximum a-posteriori prediction. The 99% confidence interval is plotted in green and based on the sample shown in figure S3.

**Figure 5:**
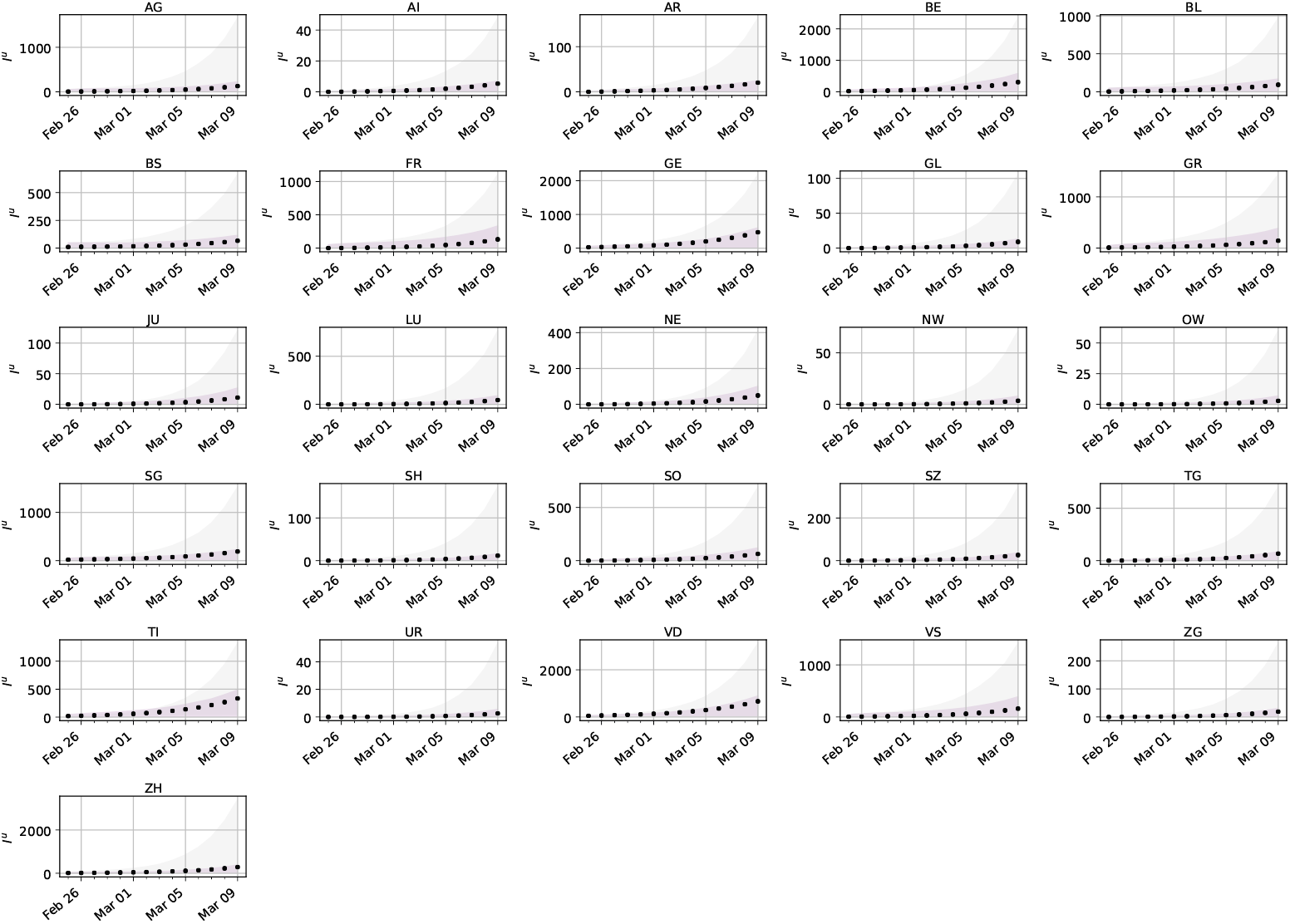
Comparison of prediction uncertainty per canton. The predictions are based on optimal strategies and non-specific testing for collection of data. They are also based on the *SEI*^*r*^*I*^*u*^*R* model output. The error bounds show the 99% confidence intervals of the unreported infectious model output for samples of the parameters with data obtained by optimal (purple) and standard testing (gray). The black dots show the actual unreported infectious for an artificial spread in Switzerland.

**Figure 6:**
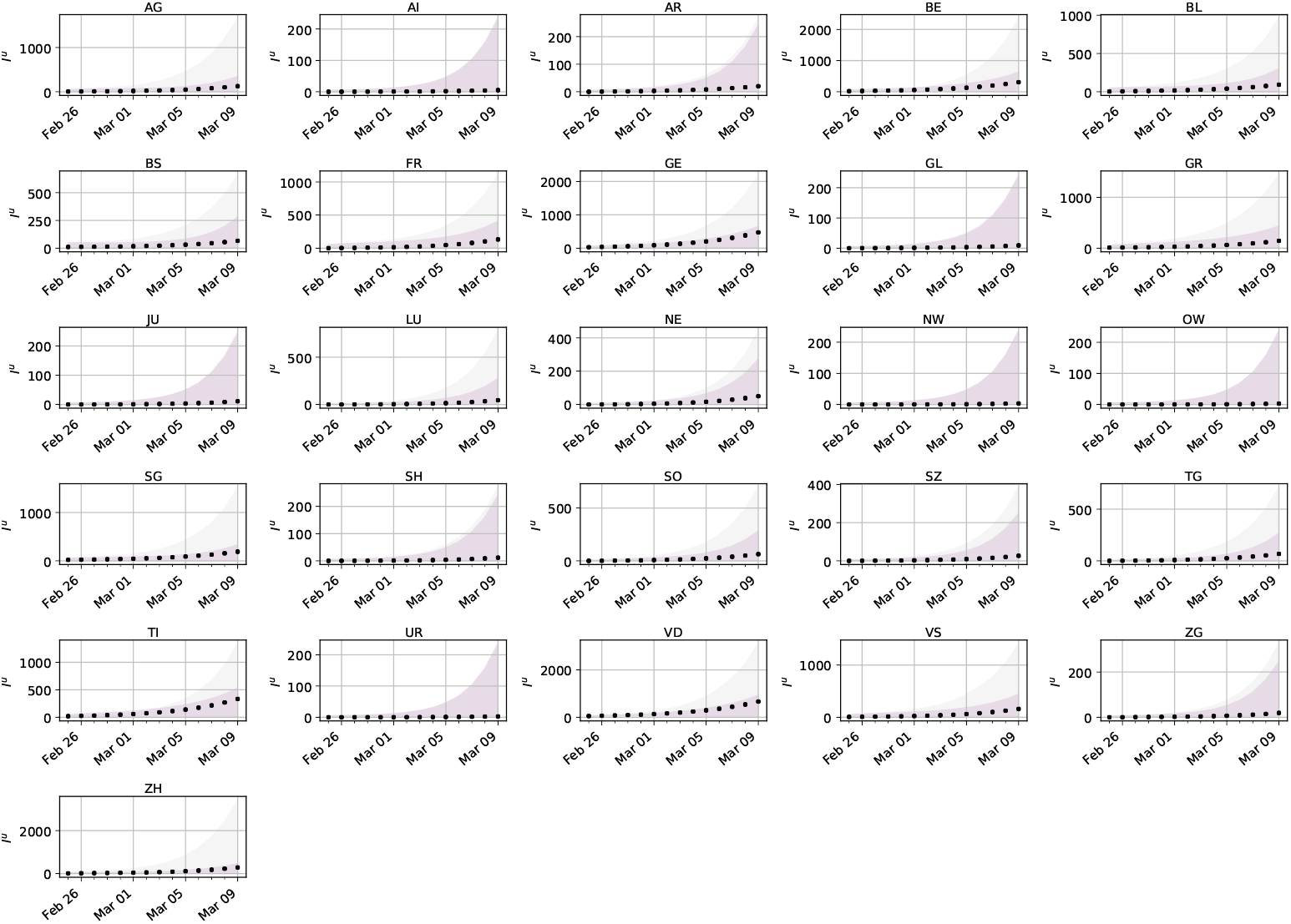
Comparison of propagated uncertainty per canton. The predictions are based on optimal strategies and non-specific testing. The *SEI*^*r*^*I*^*u*^*R* model output with added model error for the unreported infectious is shown. The error bounds show the 99% confidence intervals of the model output with added model error for samples of the parameters with data obtained by optimal (purple) and standard testing (gray). The black dots show the actual unreported infectious for an artificial spread in Switzerland.

**Table 1:** Maximum expected information gain for outbreak of a new disease. The corresponding optimal dates are shown in parenthesis.

| Canton | Maximum of Expected Information Gain |  |  |  |  |  |  |  |
| --- | --- | --- | --- | --- | --- | --- | --- | --- |
|  | 1<sup>st</sup> | measure- | 2<sup>nd</sup> | measure- | 3<sup>rd</sup> | measure- | 4<sup>th</sup> | measure- |
|  | ment |  | ment |  | ment |  | ment |  |
| AG | 2.297 | (01-03) | 1.080 | (27-02) | 0.530 | (28-02) | 0.436 | (27-02) |
| AI | 0.240 | (03-03) | 0.167 | (28-02) | 0.152 | (27-02) | 0.123 | (28-02) |
| AR | 0.538 | (03-03) | 0.189 | (28-02) | 0.157 | (03-03) | 0.127 | (26-02) |
| BE | 2.547 | (02-03) | 1.130 | (27-02) | 0.558 | (03-03) | 0.432 | (28-02) |
| BL | 2.224 | (29-02) | 1.099 | (27-02) | 0.567 | (28-02) | 0.458 | (28-02) |
| BS | 1.930 | (29-02) | 0.969 | (27-02) | 0.545 | (27-02) | 0.445 | (27-02) |
| FR | 2.046 | (01-03) | 0.983 | (27-02) | 0.533 | (28-02) | 0.424 | (27-02) |
| GE | 2.338 | (02-03) | 1.074 | (27-02) | 0.512 | (03-03) | 0.410 | (28-02) |
| GL | 0.344 | (03-03) | 0.171 | (28-02) | 0.152 | (27-02) | 0.124 | (01-03) |
| GR | 2.174 | (01-03) | 1.039 | (27-02) | 0.517 | (29-02) | 0.416 | (28-02) |
| JU | 0.408 | (03-03) | 0.176 | (29-02) | 0.154 | (01-03) | 0.125 | (03-03) |
| LU | 1.220 | (03-03) | 0.340 | (29-02) | 0.225 | (01-03) | 0.173 | (01-03) |
| NE | 0.828 | (03-03) | 0.234 | (29-02) | 0.176 | (01-03) | 0.137 | (01-03) |
| NW | 0.246 | (03-03) | 0.168 | (01-03) | 0.152 | (02-03) | 0.123 | (03-03) |
| OW | 0.225 | (03-03) | 0.168 | (03-03) | 0.152 | (02-03) | 0.124 | (03-03) |
| SG | 2.067 | (01-03) | 0.981 | (27-02) | 0.519 | (28-02) | 0.425 | (27-02) |
| SH | 0.456 | (03-03) | 0.182 | (29-02) | 0.156 | (01-03) | 0.126 | (01-03) |
| SO | 1.515 | (03-03) | 0.455 | (28-02) | 0.256 | (27-02) | 0.199 | (27-02) |
| SZ | 0.785 | (03-03) | 0.221 | (29-02) | 0.167 | (29-02) | 0.134 | (28-02) |
| TG | 1.214 | (03-03) | 0.334 | (28-02) | 0.209 | (28-02) | 0.163 | (28-02) |
| TI | 2.362 | (02-03) | 1.077 | (27-02) | 0.516 | (03-03) | 0.409 | (28-02) |
| UR | 0.210 | (03-03) | 0.167 | (03-03) | 0.152 | (02-03) | 0.124 | (28-02) |
| VD | 2.666 | (02-03) | 1.233 | (27-02) | 0.594 | (03-03) | 0.314 | (29-02) |
| VS | 2.254 | (01-03) | 1.061 | (27-02) | 0.514 | (28-02) | 0.417 | (28-02) |
| ZG | 0.701 | (03-03) | 0.206 | (28-02) | 0.161 | (29-02) | 0.130 | (28-02) |
| ZH | 2.721 | (02-03) | 1.187 | (27-02) | 0.556 | (28-02) | 0.449 | (27-02) |

**Table 2:**
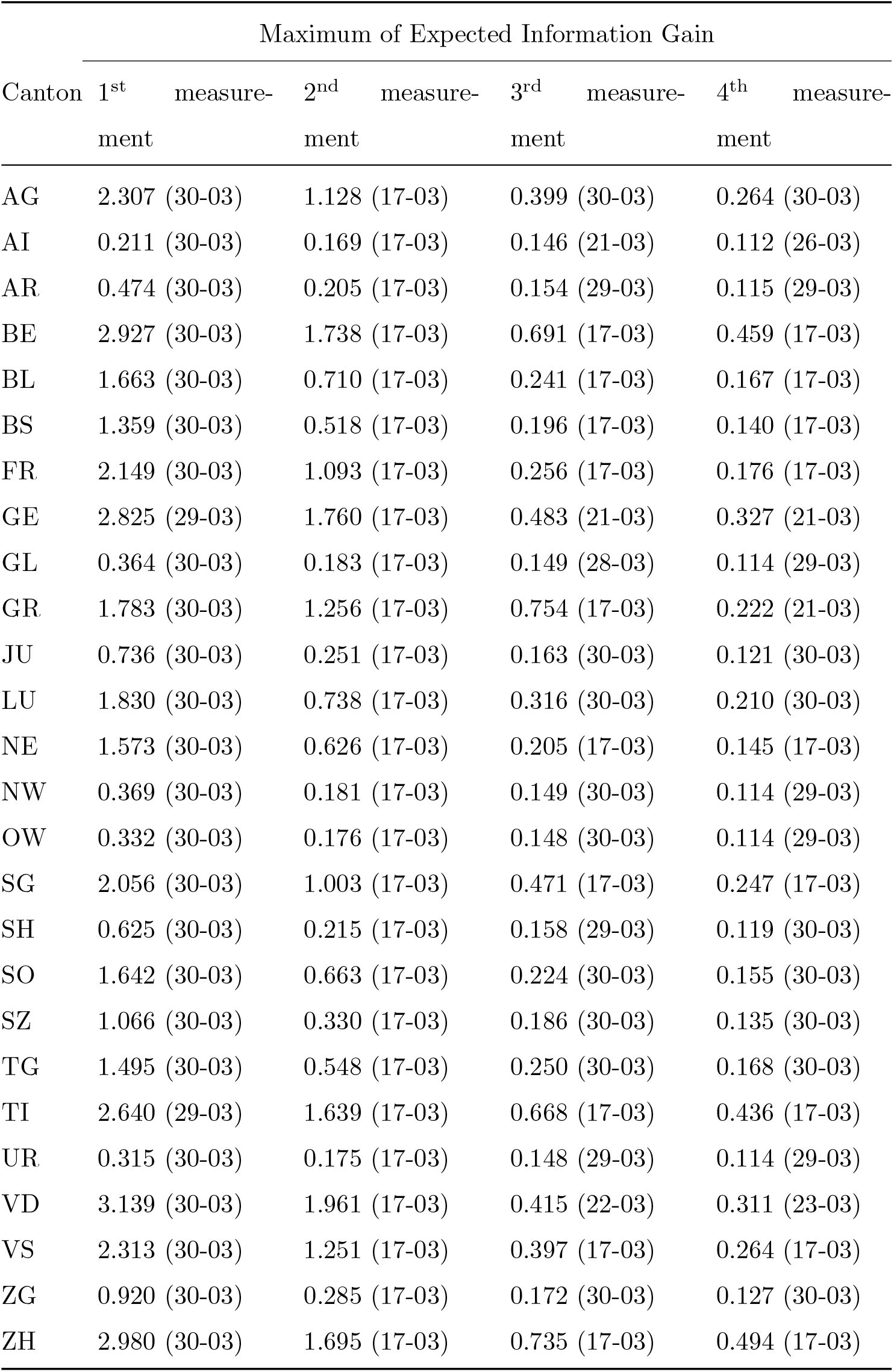
Maximum expected information gain of non-pharmaceutical interventions. The corresponding optimal dates are shown in parenthesis.

| Canton | Maximum of Expected Information Gain |  |  |  |  |  |  |  |
| --- | --- | --- | --- | --- | --- | --- | --- | --- |
|  | 1<sup>st</sup> | measure- | 2<sup>nd</sup> | measure- | 3<sup>rd</sup> | measure- | 4<sup>th</sup> | measure- |
|  | ment |  | ment |  | ment |  | ment |  |
| AG | 2.307 (30-03) |  | 1.128 (17-03) |  | 0.399 (30-03) |  | 0.264 (30-03) |  |
| AI | 0.211 (30-03) |  | 0.169 (17-03) |  | 0.146 (21-03) |  | 0.112 (26-03) |  |
| AR | 0.474 (30-03) |  | 0.205 (17-03) |  | 0.154 (29-03) |  | 0.115 (29-03) |  |
| BE | 2.927 (30-03) |  | 1.738 (17-03) |  | 0.691 (17-03) |  | 0.459 (17-03) |  |
| BL | 1.663 (30-03) |  | 0.710 (17-03) |  | 0.241 (17-03) |  | 0.167 (17-03) |  |
| BS | 1.359 (30-03) |  | 0.518 (17-03) |  | 0.196 (17-03) |  | 0.140 (17-03) |  |
| FR | 2.149 (30-03) |  | 1.093 (17-03) |  | 0.256 (17-03) |  | 0.176 (17-03) |  |
| GE | 2.825 (29-03) |  | 1.760 (17-03) |  | 0.483 (21-03) |  | 0.327 (21-03) |  |
| GL | 0.364 (30-03) |  | 0.183 (17-03) |  | 0.149 (28-03) |  | 0.114 (29-03) |  |
| GR | 1.783 (30-03) |  | 1.256 (17-03) |  | 0.754 (17-03) |  | 0.222 (21-03) |  |
| JU | 0.736 (30-03) |  | 0.251 (17-03) |  | 0.163 (30-03) |  | 0.121 (30-03) |  |
| LU | 1.830 (30-03) |  | 0.738 (17-03) |  | 0.316 (30-03) |  | 0.210 (30-03) |  |
| NE | 1.573 (30-03) |  | 0.626 (17-03) |  | 0.205 (17-03) |  | 0.145 (17-03) |  |
| NW | 0.369 (30-03) |  | 0.181 (17-03) |  | 0.149 (30-03) |  | 0.114 (29-03) |  |
| OW | 0.332 (30-03) |  | 0.176 (17-03) |  | 0.148 (30-03) |  | 0.114 (29-03) |  |
| SG | 2.056 (30-03) |  | 1.003 (17-03) |  | 0.471 (17-03) |  | 0.247 (17-03) |  |
| SH | 0.625 (30-03) |  | 0.215 (17-03) |  | 0.158 (29-03) |  | 0.119 (30-03) |  |
| SO | 1.642 (30-03) |  | 0.663 (17-03) |  | 0.224 (30-03) |  | 0.155 (30-03) |  |
| SZ | 1.066 (30-03) |  | 0.330 (17-03) |  | 0.186 (30-03) |  | 0.135 (30-03) |  |
| TG | 1.495 (30-03) |  | 0.548 (17-03) |  | 0.250 (30-03) |  | 0.168 (30-03) |  |
| TI | 2.640 (29-03) |  | 1.639 (17-03) |  | 0.668 (17-03) |  | 0.436 (17-03) |  |
| UR | 0.315 (30-03) |  | 0.175 (17-03) |  | 0.148 (29-03) |  | 0.114 (29-03) |  |
| VD | 3.139 (30-03) |  | 1.961 (17-03) |  | 0.415 (22-03) |  | 0.311 (23-03) |  |
| VS | 2.313 (30-03) |  | 1.251 (17-03) |  | 0.397 (17-03) |  | 0.264 (17-03) |  |
| ZG | 0.920 (30-03) |  | 0.285 (17-03) |  | 0.172 (30-03) |  | 0.127 (30-03) |  |
| ZH | 2.980 (30-03) |  | 1.695 (17-03) |  | 0.735 (17-03) |  | 0.494 (17-03) |  |

**Table 3:** Maximum expected information gain for monitoring of a second outbreak with uninformed *b*_3_. The corresponding optimal dates are shown in parenthesis.

| Canton | Maximum of Expected Information Gain |  |  |  |  |  |  |  |
| --- | --- | --- | --- | --- | --- | --- | --- | --- |
|  | 1<sup>st</sup> | measure- | 2<sup>nd</sup> | measure- | 3<sup>rd</sup> | measure- | 4<sup>th</sup> | measure- |
|  | ment |  | ment |  | ment |  | ment |  |
| AG | 1.356 | (13-06) | 0.434 | (06-06) | 0.292 | (06-06) | 0.210 | (13-06) |
| AI | 0.171 | (13-06) | 0.167 | (09-06) | 0.159 | (09-06) | 0.142 | (08-06) |
| AR | 0.207 | (13-06) | 0.169 | (08-06) | 0.159 | (08-06) | 0.142 | (11-06) |
| BE | 1.877 | (13-06) | 0.746 | (06-06) | 0.435 | (06-06) | 0.339 | (13-06) |
| BL | 0.712 | (13-06) | 0.236 | (07-06) | 0.186 | (06-06) | 0.156 | (13-06) |
| BS | 0.512 | (13-06) | 0.202 | (06-06) | 0.172 | (06-06) | 0.148 | (13-06) |
| FR | 0.973 | (13-06) | 0.330 | (06-06) | 0.208 | (13-06) | 0.184 | (13-06) |
| GE | 1.490 | (13-06) | 0.644 | (06-06) | 0.381 | (13-06) | 0.328 | (13-06) |
| GL | 0.189 | (13-06) | 0.168 | (08-06) | 0.159 | (12-06) | 0.142 | (08-06) |
| GR | 0.567 | (13-06) | 0.219 | (06-06) | 0.173 | (06-06) | 0.152 | (13-06) |
| JU | 0.255 | (13-06) | 0.173 | (07-06) | 0.161 | (06-06) | 0.143 | (13-06) |
| LU | 0.936 | (13-06) | 0.286 | (07-06) | 0.213 | (06-06) | 0.168 | (13-06) |
| NE | 0.576 | (13-06) | 0.222 | (06-06) | 0.172 | (13-06) | 0.153 | (13-06) |
| NW | 0.193 | (13-06) | 0.168 | (07-06) | 0.159 | (06-06) | 0.142 | (08-06) |
| OW | 0.187 | (13-06) | 0.167 | (06-06) | 0.159 | (07-06) | 0.142 | (11-06) |
| SG | 1.084 | (13-06) | 0.330 | (06-06) | 0.238 | (06-06) | 0.179 | (13-06) |
| SH | 0.247 | (13-06) | 0.172 | (07-06) | 0.161 | (06-06) | 0.142 | (07-06) |
| SO | 0.701 | (13-06) | 0.235 | (06-06) | 0.184 | (06-06) | 0.154 | (13-06) |
| SZ | 0.403 | (13-06) | 0.187 | (07-06) | 0.167 | (06-06) | 0.145 | (13-06) |
| TG | 0.651 | (13-06) | 0.223 | (06-06) | 0.183 | (06-06) | 0.153 | (13-06) |
| TI | 1.241 | (13-06) | 0.539 | (06-06) | 0.367 | (13-06) | 0.322 | (13-06) |
| UR | 0.186 | (13-06) | 0.167 | (09-06) | 0.158 | (09-06) | 0.142 | (08-06) |
| VD | 1.881 | (13-06) | 0.873 | (06-06) | 0.505 | (13-06) | 0.443 | (13-06) |
| VS | 1.138 | (13-06) | 0.420 | (06-06) | 0.256 | (13-06) | 0.223 | (13-06) |
| ZG | 0.337 | (13-06) | 0.180 | (07-06) | 0.163 | (07-06) | 0.144 | (13-06) |
| ZH | 2.092 | (13-06) | 0.862 | (06-06) | 0.592 | (06-06) | 0.276 | (09-06) |

Assume we want to estimate the proportion of a population with some margin of error *d* and a small risk *α*, i.e., we want Pr(|*P* − *p*| ≥ *d*) = *α*. Here, the proportion corresponds to the proportion of unreported infected population. The minimum number of samples to achieve this is given by Cochran’s formula [14],

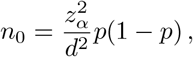

where *z*_*α*_ is the inverse of the standard normal cumulative distribution function evaluated at 1 − *α/*2. In this formula, we have assumed that the population is of infinite size. In order to correct for a finite size population *N*, we compute

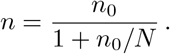

In the next figure we present the minimum number of samples needed to sample the cantons of Switzerland for *d* = 0.01 and *α* = 0.01. Notice that *α* = 0.01 corresponds to a 99% confidence interval.

If the available test-kits are more than 26×5950 = 154700 then the maximum information gain will be achieved by deploying all tests uniformly in all cantons. However, when it is not realistic to conduct over 154700 tests, we consider testing with limited resources. For example assuming 30000 available tests, will be enough to test 5 cantons 5 × 5950. The question we answer then is which 5 cantons (from the 26) should we test given that we must test a minimum population of 5950 per canton?.

Distributing less than a particular number of tests (5950) in a canton will not provide a statistically reliable estimate for the number of unreported infections there. Thus, in such a case, the measured unreported infections should not be used to estimate the expected information gain.

Finally, we note that in this work we ignore the bias in the estimate of *I*^*u*^. This means that the estimates of unreported infected enter the Bayesian framework without explicitly accounting for this known error.

**Table 4:** Maximum expected information gain to monitor a second outbreak with informed *b*_3_. The corresponding optimal dates are shown in parenthesis.

| Canton | Maximum of Expected Information Gain |  |  |  |  |  |  |  |
| --- | --- | --- | --- | --- | --- | --- | --- | --- |
|  | 1<sup>st</sup><br>ment | measure-<br>ment | 2<sup>nd</sup><br>ment | measure-<br>ment | 3<sup>rd</sup><br>ment | measure-<br>ment | 4<sup>th</sup><br>ment | measure-<br>ment |
| AG | 1.233 (17-07) |  | 0.328 (10-07) |  | 0.220 (17-07) |  | 0.194 (10-07) |  |
| AI | 0.170 (17-07) |  | 0.167 (17-07) |  | 0.154 (13-07) |  | 0.136 (17-07) |  |
| AR | 0.198 (17-07) |  | 0.169 (10-07) |  | 0.154 (14-07) |  | 0.136 (16-07) |  |
| BE | 1.596 (17-07) |  | 0.460 (10-07) |  | 0.347 (17-07) |  | 0.267 (10-07) |  |
| BL | 0.616 (17-07) |  | 0.202 (10-07) |  | 0.166 (17-07) |  | 0.147 (10-07) |  |
| BS | 0.441 (17-07) |  | 0.184 (10-07) |  | 0.159 (17-07) |  | 0.141 (10-07) |  |
| FR | 0.636 (17-07) |  | 0.209 (10-07) |  | 0.193 (17-07) |  | 0.156 (17-07) |  |
| GE | 0.896 (17-07) |  | 0.326 (17-07) |  | 0.308 (17-07) |  | 0.225 (17-07) |  |
| GL | 0.184 (17-07) |  | 0.168 (13-07) |  | 0.154 (17-07) |  | 0.136 (10-07) |  |
| GR | 0.418 (17-07) |  | 0.182 (10-07) |  | 0.162 (17-07) |  | 0.141 (10-07) |  |
| JU | 0.219 (17-07) |  | 0.169 (10-07) |  | 0.155 (17-07) |  | 0.136 (17-07) |  |
| LU | 0.834 (17-07) |  | 0.234 (10-07) |  | 0.178 (17-07) |  | 0.157 (10-07) |  |
| NE | 0.378 (17-07) |  | 0.180 (10-07) |  | 0.165 (17-07) |  | 0.142 (17-07) |  |
| NW | 0.187 (17-07) |  | 0.168 (10-07) |  | 0.154 (15-07) |  | 0.135 (10-07) |  |
| OW | 0.183 (17-07) |  | 0.168 (10-07) |  | 0.154 (10-07) |  | 0.136 (12-07) |  |
| SG | 0.994 (17-07) |  | 0.267 (10-07) |  | 0.191 (17-07) |  | 0.169 (10-07) |  |
| SH | 0.232 (17-07) |  | 0.170 (10-07) |  | 0.154 (15-07) |  | 0.136 (11-07) |  |
| SO | 0.581 (17-07) |  | 0.197 (10-07) |  | 0.166 (17-07) |  | 0.145 (10-07) |  |
| SZ | 0.362 (17-07) |  | 0.178 (10-07) |  | 0.157 (10-07) |  | 0.139 (10-07) |  |
| TG | 0.591 (17-07) |  | 0.200 (10-07) |  | 0.164 (17-07) |  | 0.145 (10-07) |  |
| TI | 0.556 (17-07) |  | 0.318 (17-07) |  | 0.296 (17-07) |  | 0.224 (17-07) |  |
| UR | 0.181 (17-07) |  | 0.168 (11-07) |  | 0.154 (16-07) |  | 0.135 (16-07) |  |
| VD | 1.297 (17-07) |  | 0.452 (17-07) |  | 0.433 (17-07) |  | 0.281 (10-07) |  |
| VS | 0.655 (17-07) |  | 0.245 (17-07) |  | 0.228 (17-07) |  | 0.176 (17-07) |  |
| ZG | 0.303 (17-07) |  | 0.174 (10-07) |  | 0.155 (15-07) |  | 0.137 (11-07) |  |
| ZH | 1.976 (17-07) |  | 0.675 (10-07) |  | 0.276 (14-07) |  | 0.250 (13-07) |  |

**Table 5:** Parameters and prior distributions used in Bayesian inference. Here the data corresponds to the daily reported infections. In all cases, data are used from the 25^th^ of February 2020, when the first reported case was found in the canton of Ticino. Inference I uses data up to the day non-pharmaceutical interventions were announced (17^th^ of March 2020). Inference II uses data up to the day measures were relaxed (6^th^ of June 2020). Inference III uses data up to the 9^th^ of July 2020. The choice of prior distributions is consistent with the choice found in [9]; the ranges used in our study are slightly extended.

| Parameter | Prior distribution / fixed value | Inference I | Inference II | Inference III |
| --- | --- | --- | --- | --- |
| $b_0$ | $\mathcal{U}([0.8, 1.8])$ | yes | yes | yes |
| $\alpha$ | $\mathcal{U}([0.01, 1])$ | yes | yes | yes |
| $\mu$ | $\mathcal{U}([0.2, 1])$ | yes | yes | yes |
| $Z$ | $\mathcal{U}([1, 6])$ | yes | yes | yes |
| $D$ | $\mathcal{U}([1, 6])$ | yes | yes | yes |
| $\theta_0$ | $\mathcal{U}([0.5, 1.5])$ | yes | yes | yes |
| $c_1$ | $\mathcal{U}([0, 1])$ | no | yes | yes |
| $c_2$ | $\mathcal{U}([0, 1])$ | no | yes | yes |
| $c_3$ | $\mathcal{U}([0, 1])$ | no | yes | yes |
| $c_4$ | $\mathcal{U}([0, 1])$ | no | yes | yes |
| $\delta_1$ | $\mathcal{U}([20, 30])$ | no | yes | yes |
| $\delta_2$ | $\mathcal{U}([30, 40])$ | no | yes | yes |
| $\delta_3$ | 102 | no | yes | yes |
| $\lambda$ | $\mathcal{U}([0, 0.03])$ | no | no | yes |
| $r$ | $\mathcal{U}([0, 2])$ | yes | yes | yes |
| $\mathbf{I}_{\text{IC}}^u$ | $\mathcal{U}([0, 50]^{12})$ | yes | yes | yes |

**Figure 7:**
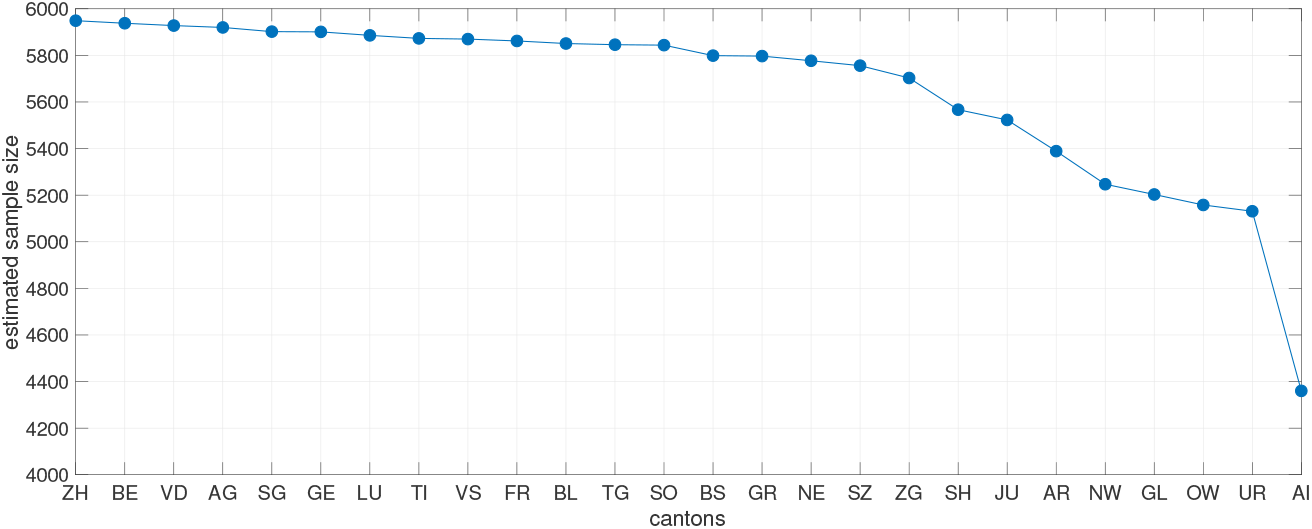
Estimated sample size using Cochran’s [14] formula for every canton for confidence level 99%, margin of error 1% and probability of infection 0.1. The cantons are sorted in descending order of their population. The maximum sample size is estimated for Zurich and is equal to 5950. All the other cantons need up to 14% less samples with the exception of the smallest canton that needs 27% less samples.

